# Red blood cell distribution width-derived indces predicts long-term prognosis in acute heart failure

**DOI:** 10.1101/2024.12.22.24319514

**Authors:** Miao Zhang, Jing Zhu, Degang Mo, Shanshan Yuan, Fanhui Lin, Hongyan Dai

## Abstract

**Background:** Red blood cell distribution width (RDW), a routine indicator of erythrocyte volume variability, has shown potential in recent years in the prognostic assessment of a variety of diseases, including acute heart failure (AHF). The predictive ability of RDW-derived indices, namely the hemoglobin-to-RDW ratio (HRR), the RDW-to-lymphocyte ratio (RLR), the RDW-to-platelet ratio (RPR), the RDW-to-albumin ratio (RAR), and the product of RDW and mean corpuscular volume (RDW×MCV), for the prognosis of AHF remains unclear.

**Methods:** The study included 4090 eligible patients in the MIMIC 3.0 database, screened variables using Lasso regression, assessed whether these derivatives independently predicted 1-year mortality from AHF by Cox proportional hazards model, and plotted survival curves and time-related ROC curves. Finally, the relationship between each indicator and outcome was analyzed by restricted cubic spline.

**Results:** Positive events occurred in 2085 (51%) patients with decreased HRR and increased RLR, RPR, RAR, and RDW×MCV (P<0.001).After Cox model adjustment, HRR, RPR, RAR, and RDW×MCV remained independent predictors of 1-year prognosis for AHF (RLR was not) and the relationship may be more linear. Of these, RPR had the greatest impact on survival time (HR = 1.89, 95%CI: 1.33-2.67) and RDW×MCV had the highest predictive efficacy (AUC=0.612).

**Conclusion:** RDW-derived indices HRR, RPR, RAR and RDW×MCV independently predicted 1-year mortality in AHF, and RLR had no independent predictive value.

## 1 Introduction

Acute Heart Failure (AHF), a serious disease in the cardiovascular field, with its high morbidity and mortality has been a major challenge in global public health(1).Although the treatment of chronic HF continues to receive the good news of new drug approvals, in the field of AHF, there have been few new therapies to improve mortality for decades (2), which has led to its high mortality rate, which has been estimated to be as high as about 33% at 1 year (1). “Time is prognosis” and any delay in the initiation of therapy is associated with poorer outcomes(3). Therefore, the search for validated prognostic predictors to optimize AHF therapy and develop follow-up methods in the context of being able to identify high-risk patients as early as possible is a useful, efficient, and cost-effective way to reduce the economic burden of AHF and improve outcomes(3).

As an important index reflecting the heterogeneity of erythrocyte volume in peripheral blood, red blood cell distribution width (RDW) has gradually gained wide attention in the field of cardiovascular diseases in recent years(4–9). Several studies have demonstrated the prognostic value of RDW in AHF. Van Kimmenade et al. showed that RDW was increased in patients with AHF and independently predicted their 1-year mortality(10); Makhoul et al. also demonstrated that RDW was a strong independent predictor of higher morbidity and 1-year mortality(11); Uemura et al. showed that during hospitalization changes in RDW values, but not RDW values at admission, independently predicted adverse outcomes in acute decompensated HF (ADHF) (12); and Zhang et al. showed that increased RDW during hospitalization was independently associated with short- or long-term all-cause mortality in critically ill patients with HF, including AHF cohort(13). Together, these studies establish the prognostic value of RDW in AHF.

However, Poz et al. suggested that the pathophysiology of cardiovascular disease is extremely complex and that a single monitoring of erythrocytes does not represent the most accurate strategy for predicting cardiac disease(5), while in recent years several studies have begun to investigate the prognostic impact of RDW-derived indices on the disease(14–18). Therefore, in this paper, RDW was combined with other indices, respectively, i.e., hemoglobin (Hb) to RDW ratio (HRR), RDW to absolute lymphocyte count (ALC) ratio (RLR), RDW to platelet count (PLT) ratio (RPR), RDW to albumin ratio (RAR), and the product of RDW and mean corpuscular volume (RDW × MCV), to explore whether these derived indices still have an independent predictive value of 1-year mortality rate of patients with AHF, which can help physicians to more accurately assess the patient’s condition and risk, and to formulate more reasonable treatment program.

## 2 Methods

### 2.1 Study population collection

We used data derived from the MIMIC-IV 3.0 database(19), a contemporary, publicly available electronic health record dataset on intensive care units. The dataset covers twelve years of admissions at Beth Israel Deaconess Medical Center from 2008 to 2022, and contains a wealth of information about clinical care. We successfully extracted the required data by utilizing custom SQL scripts, converting the data to MIMIC standard structure, and executing SQL query statements through Navicat Premium 16 software. De-identification measures were taken to protect patient privacy, so it was not necessary to obtain informed consent from the patients. The author, Degang Mo (ID: 65748833), has been authorized to access the database after completing the relevant training.

### 2.2 Inclusion and exclusion criteria of study subjects

We initially screened 9909 patients who were older than 18 years of age and were first admitted to the ICU with an ICD code-confirmed diagnosis of AHF from the MIMIC-IV 3.0 database. Subsequently, 5788 individuals missing key information such as RDW, Hb, ALC, PLT, MCV, and albumin were excluded, as well as 31 patients who were admitted for less than 24 hours. Ultimately, 4090 patients were included in the study cohort (see Figure 1).

**Figure 1.**
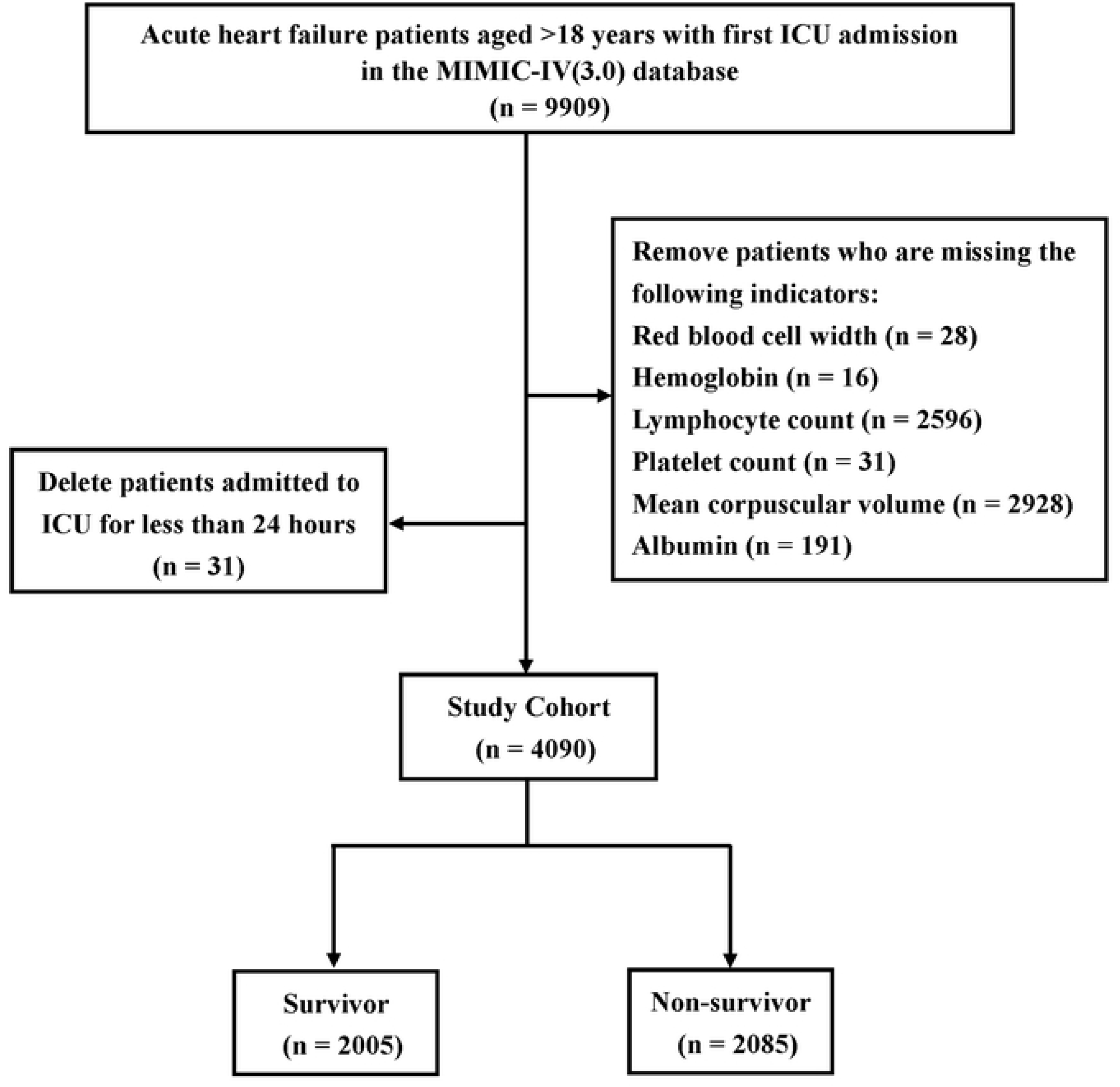
Flowchart of the study population.

### 2.3 Baseline characteristics of the study population

As shown in Table 1, we collected the following information: demographic data, including age, gender, weight, urine output on the first day of admission to the ICU, as well as physiologic indices such as average heart rate (HR), respiratory rate, systolic blood pressure (SBP), diastolic blood pressure, body temperature and arterial oxygen saturation on the first day. Laboratory test data covered erythrocyte, neutrophil, ALC, monocyte, eosinophil, basophil, and PLT, as well as Hb, MCV, erythrocyte distribution width, albumin, glutamatergic aminotransferase, serum creatinine, NT-proBNP, plasminogen time, lactate, blood sodium, partial pressure of carbon dioxide in blood gas analysis, and PH values. For comorbidities, we recorded myocardial infarction, peripheral vascular disease, cerebrovascular disease, dementia, chronic lung disease, rheumatism, peptic ulcer, mild fatty liver disease, diabetes mellitus (with or without comorbidities), paraplegia, nephropathy, malignant tumors, severe liver disease, and metastatic solid tumors. Post-admission drug use, on the other hand, included angiotensin-converting enzyme inhibitors (ACEI), angiotensin II receptor blockers, β-blockers, calcium channel blockers (CCB), diuretics, nitroglycerin, simvastatin, antiplatelet agents, anticoagulants, amiodarone, digoxin and sodium bicarbonate. In addition, we included SOFA scores to assess disease severity. To improve data quality, we excluded laboratory indicators with more than 20% missing values, filled in the median of the missing values using Stata MP 17, and identified and replaced outliers using the “capping method”.

### 2.4 Calculation of RDW-derived indices

HRR = Hb (g/L) / RDW (%)

RLR = RDW (%) / ALC (×10^9^/L)

RPR = RDW (%) / PLT (×10^9^/L)

RAR = RDW (%) / albumin (g/dL)

RDW x MCV = RDW (%) × MCV (fL)/100

### 2.5 Endpoints

The final outcome of the study is concerned with the survival status of the patient at 1 year, i.e., determining whether the patient has died.

### 2.6 Statistics

The Shapiro-Wilk test was used to determine whether the numerical variables were normally distributed. For data that did not conform to normal distribution, we used the median (and interquartile range) to characterize them, and the Mann-Whitney U test to compare whether there were statistically significant differences between the two groups. For categorical variables, we present them in terms of their frequencies (and percentages) and apply the chi-square test to compare between groups. Least Absolute Shrinkage and Selection Operator (Lasso) regression were used to perform variable screening for each RDW-related derived indicator, respectively, and 10-fold cross-validation was used to determine the optimal regularization parameter, λ-1se, and to fit the model via the glmnet package. The screened variables were included in univariate and multivariate Cox proportional hazards regression model (Cox Model) to determine the independent factors associated with 1-year mortality in AHF, respectively. Variables with p < 0.05 in the univariate Cox Model were included in the multivariate Cox Model, and variables were screened using the backward method to determine whether each RDW-derived index was an independent predictor of poor prognosis in AHF in this study. The cut-off values of the RDW-derived indicators with independent predictive value screened by appeal were calculated using Jamovi software, thus categorizing the indicators into two groups, while survival curves were plotted using the Kaplan-Meier method,and time-ROC curves were plotted. Finally, restricted cubic spline (RCS) was used to explore whether there was a nonlinear association between the appeal continuum variables and the outcome. Data were processed and analyzed using R (version 4.0.2) and SPSS (version 25). A two-sided p < 0.05 was considered statistically significant.

## 3 Results

### 3.1 Compare baseline characteristics of AHF patients with different outcomes

We successfully enrolled 4090 patients into the study cohort and categorized them into a survival group (2005,49%) and a death group (2085,51%) based on their survival status at 1 year. Comparing the data of the two groups, we found that the patients in the death group were generally characterized by older age, lower body weight, accelerated HR, decreased blood pressure, hypothermia and decreased urine output at the time of admission, and these differences were statistically significant. In terms of laboratory tests, patients in the mortality group had low red blood cell, Hb, ALC, eosinophil, basophil, and PLT, as well as relatively low albumin and pH values, while the MCV, RDW, NT-proBNP, lactate content, and partial pressure of carbon dioxide were high, and the prothrombin time was prolonged. In addition, in terms of RDW-derived indices, patients who experienced adverse outcomes exhibited higher RLR, RPR, RAR, and RDW × MCV values on admission, whereas HRR was relatively low, and the difference was statistically significant. In terms of comorbidities, patients who died were more frequently accompanied by peripheral vascular disease, cerebrovascular disease, dementia, chronic pulmonary disease, diabetes mellitus with co-morbidities, renal disease, Malignant Cancer, severe liver disease, metastatic solid tumor. In terms of medication history, patients in the death group used ACEI, nitroglycerin, and anticoagulants less frequently after admission, whereas they used CCB, β -blockers, simvastatin, amiodarone, digoxin, and sodium bicarbonate more frequently. In terms of scoring, patients in the death group had significantly higher post-hospitalization Sofa scores than those in the survival group (see Table 1).

**Table 1.**
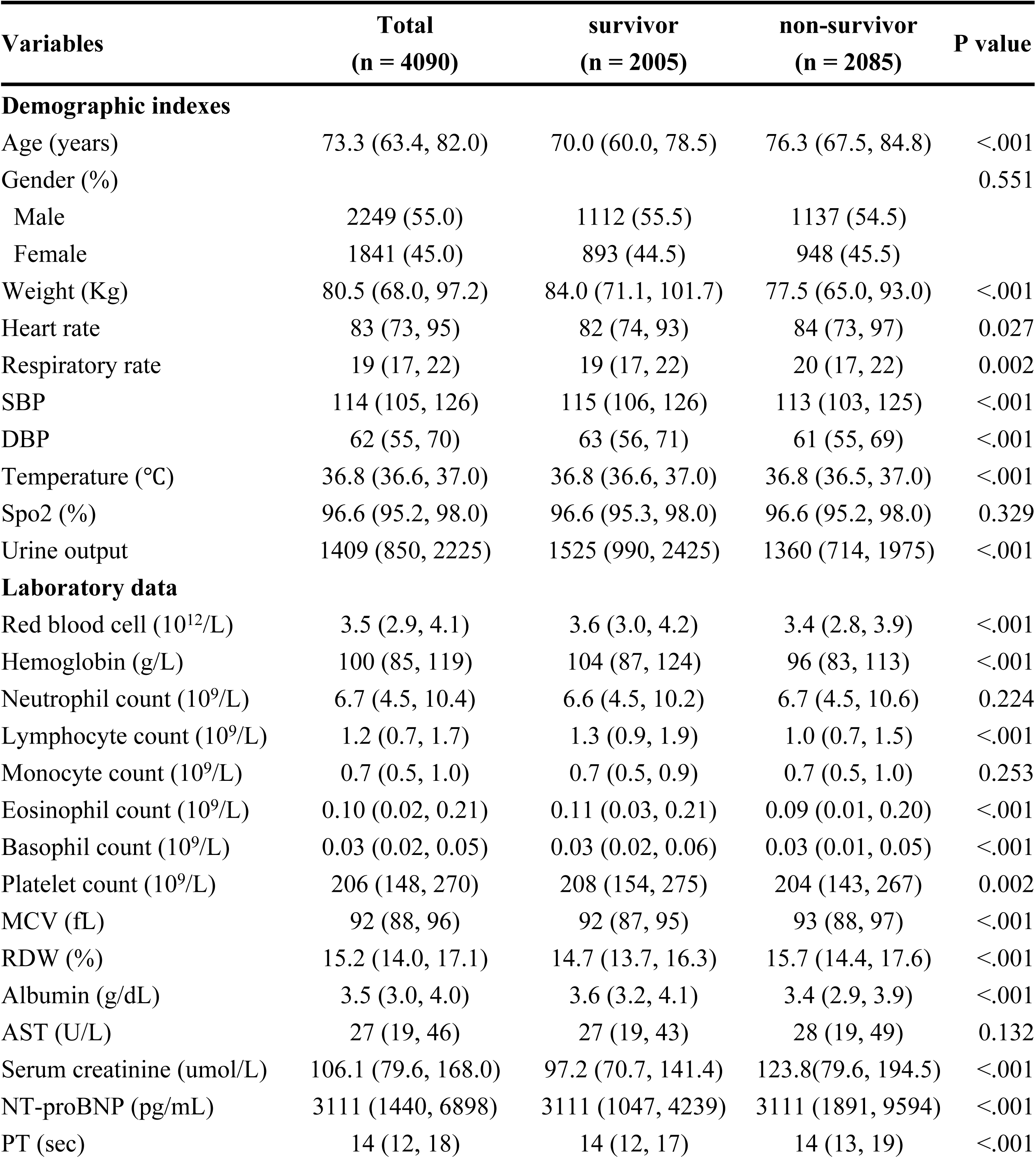
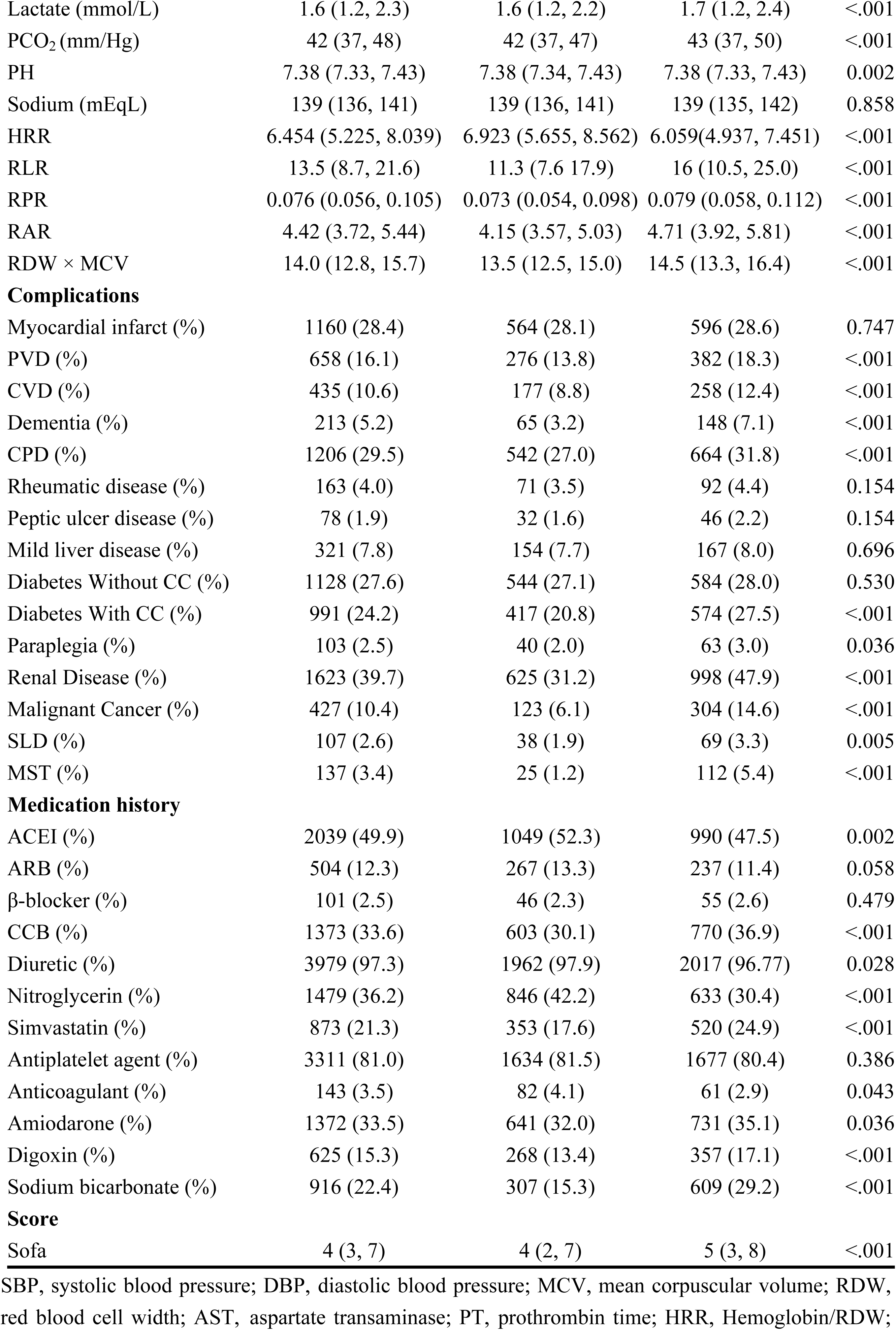
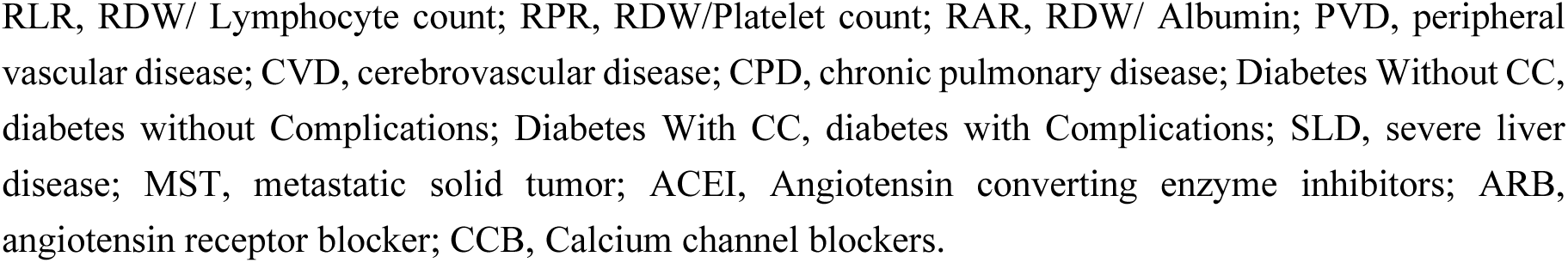
Compare baseline characteristics of AHF patients with different outcomes.

| Variables | Total<br>(n = 4090) | survivor<br>(n = 2005) | non-survivor<br>(n = 2085) | P value |
| --- | --- | --- | --- | --- |
| **Demographic indexes** |  |  |  |  |
| Age (years) | 73.3 (63.4, 82.0) | 70.0 (60.0, 78.5) | 76.3 (67.5, 84.8) | <.001 |
| Gender (%) |  |  |  | 0.551 |
| Male | 2249 (55.0) | 1112 (55.5) | 1137 (54.5) |  |
| Female | 1841 (45.0) | 893 (44.5) | 948 (45.5) |  |
| Weight (Kg) | 80.5 (68.0, 97.2) | 84.0 (71.1, 101.7) | 77.5 (65.0, 93.0) | <.001 |
| Heart rate | 83 (73, 95) | 82 (74, 93) | 84 (73, 97) | 0.027 |
| Respiratory rate | 19 (17, 22) | 19 (17, 22) | 20 (17, 22) | 0.002 |
| SBP | 114 (105, 126) | 115 (106, 126) | 113 (103, 125) | <.001 |
| DBP | 62 (55, 70) | 63 (56, 71) | 61 (55, 69) | <.001 |
| Temperature (°C) | 36.8 (36.6, 37.0) | 36.8 (36.6, 37.0) | 36.8 (36.5, 37.0) | <.001 |
| Spo2 (%) | 96.6 (95.2, 98.0) | 96.6 (95.3, 98.0) | 96.6 (95.2, 98.0) | 0.329 |
| Urine output | 1409 (850, 2225) | 1525 (990, 2425) | 1360 (714, 1975) | <.001 |
| **Laboratory data** |  |  |  |  |
| Red blood cell (10<sup>12</sup>/L) | 3.5 (2.9, 4.1) | 3.6 (3.0, 4.2) | 3.4 (2.8, 3.9) | <.001 |
| Hemoglobin (g/L) | 100 (85, 119) | 104 (87, 124) | 96 (83, 113) | <.001 |
| Neutrophil count (10<sup>9</sup>/L) | 6.7 (4.5, 10.4) | 6.6 (4.5, 10.2) | 6.7 (4.5, 10.6) | 0.224 |
| Lymphocyte count (10<sup>9</sup>/L) | 1.2 (0.7, 1.7) | 1.3 (0.9, 1.9) | 1.0 (0.7, 1.5) | <.001 |
| Monocyte count (10<sup>9</sup>/L) | 0.7 (0.5, 1.0) | 0.7 (0.5, 0.9) | 0.7 (0.5, 1.0) | 0.253 |
| Eosinophil count (10<sup>9</sup>/L) | 0.10 (0.02, 0.21) | 0.11 (0.03, 0.21) | 0.09 (0.01, 0.20) | <.001 |
| Basophil count (10<sup>9</sup>/L) | 0.03 (0.02, 0.05) | 0.03 (0.02, 0.06) | 0.03 (0.01, 0.05) | <.001 |
| Platelet count (10<sup>9</sup>/L) | 206 (148, 270) | 208 (154, 275) | 204 (143, 267) | 0.002 |
| MCV (fL) | 92 (88, 96) | 92 (87, 95) | 93 (88, 97) | <.001 |
| RDW (%) | 15.2 (14.0, 17.1) | 14.7 (13.7, 16.3) | 15.7 (14.4, 17.6) | <.001 |
| Albumin (g/dL) | 3.5 (3.0, 4.0) | 3.6 (3.2, 4.1) | 3.4 (2.9, 3.9) | <.001 |
| AST (U/L) | 27 (19, 46) | 27 (19, 43) | 28 (19, 49) | 0.132 |
| Serum creatinine (umol/L) | 106.1 (79.6, 168.0) | 97.2 (70.7, 141.4) | 123.8(79.6, 194.5) | <.001 |
| NT-proBNP (pg/mL) | 3111 (1440, 6898) | 3111 (1047, 4239) | 3111 (1891, 9594) | <.001 |
| PT (sec) | 14 (12, 18) | 14 (12, 17) | 14 (13, 19) | <.001 |
| Lactate (mmol/L) | 1.6 (1.2, 2.3) | 1.6 (1.2, 2.2) | 1.7 (1.2, 2.4) | <.001 |
| PCO<sub>2</sub> (mm/Hg) | 42 (37, 48) | 42 (37, 47) | 43 (37, 50) | <.001 |
| PH | 7.38 (7.33, 7.43) | 7.38 (7.34, 7.43) | 7.38 (7.33, 7.43) | 0.002 |
| Sodium (mEq/L) | 139 (136, 141) | 139 (136, 141) | 139 (135, 142) | 0.858 |
| HRR | 6.454 (5.225, 8.039) | 6.923 (5.655, 8.562) | 6.059(4.937, 7.451) | <.001 |
| RLR | 13.5 (8.7, 21.6) | 11.3 (7.6 17.9) | 16 (10.5, 25.0) | <.001 |
| RPR | 0.076 (0.056, 0.105) | 0.073 (0.054, 0.098) | 0.079 (0.058, 0.112) | <.001 |
| RAR | 4.42 (3.72, 5.44) | 4.15 (3.57, 5.03) | 4.71 (3.92, 5.81) | <.001 |
| RDW × MCV | 14.0 (12.8, 15.7) | 13.5 (12.5, 15.0) | 14.5 (13.3, 16.4) | <.001 |
| **Complications** |  |  |  |  |
| Myocardial infarct (%) | 1160 (28.4) | 564 (28.1) | 596 (28.6) | 0.747 |
| PVD (%) | 658 (16.1) | 276 (13.8) | 382 (18.3) | <.001 |
| CVD (%) | 435 (10.6) | 177 (8.8) | 258 (12.4) | <.001 |
| Dementia (%) | 213 (5.2) | 65 (3.2) | 148 (7.1) | <.001 |
| CPD (%) | 1206 (29.5) | 542 (27.0) | 664 (31.8) | <.001 |
| Rheumatic disease (%) | 163 (4.0) | 71 (3.5) | 92 (4.4) | 0.154 |
| Peptic ulcer disease (%) | 78 (1.9) | 32 (1.6) | 46 (2.2) | 0.154 |
| Mild liver disease (%) | 321 (7.8) | 154 (7.7) | 167 (8.0) | 0.696 |
| Diabetes Without CC (%) | 1128 (27.6) | 544 (27.1) | 584 (28.0) | 0.530 |
| Diabetes With CC (%) | 991 (24.2) | 417 (20.8) | 574 (27.5) | <.001 |
| Paraplegia (%) | 103 (2.5) | 40 (2.0) | 63 (3.0) | 0.036 |
| Renal Disease (%) | 1623 (39.7) | 625 (31.2) | 998 (47.9) | <.001 |
| Malignant Cancer (%) | 427 (10.4) | 123 (6.1) | 304 (14.6) | <.001 |
| SLD (%) | 107 (2.6) | 38 (1.9) | 69 (3.3) | 0.005 |
| MST (%) | 137 (3.4) | 25 (1.2) | 112 (5.4) | <.001 |
| **Medication history** |  |  |  |  |
| ACEI (%) | 2039 (49.9) | 1049 (52.3) | 990 (47.5) | 0.002 |
| ARB (%) | 504 (12.3) | 267 (13.3) | 237 (11.4) | 0.058 |
| β-blocker (%) | 101 (2.5) | 46 (2.3) | 55 (2.6) | 0.479 |
| CCB (%) | 1373 (33.6) | 603 (30.1) | 770 (36.9) | <.001 |
| Diuretic (%) | 3979 (97.3) | 1962 (97.9) | 2017 (96.77) | 0.028 |
| Nitroglycerin (%) | 1479 (36.2) | 846 (42.2) | 633 (30.4) | <.001 |
| Simvastatin (%) | 873 (21.3) | 353 (17.6) | 520 (24.9) | <.001 |
| Antiplatelet agent (%) | 3311 (81.0) | 1634 (81.5) | 1677 (80.4) | 0.386 |
| Anticoagulant (%) | 143 (3.5) | 82 (4.1) | 61 (2.9) | 0.043 |
| Amiodarone (%) | 1372 (33.5) | 641 (32.0) | 731 (35.1) | 0.036 |
| Digoxin (%) | 625 (15.3) | 268 (13.4) | 357 (17.1) | <.001 |
| Sodium bicarbonate (%) | 916 (22.4) | 307 (15.3) | 609 (29.2) | <.001 |
| **Score** |  |  |  |  |
| Sofa | 4 (3, 7) | 4 (2, 7) | 5 (3, 8) | <.001 |

SBP, systolic blood pressure; DBP, diastolic blood pressure; MCV, mean corpuscular volume; RDW, red blood cell width; AST, aspartate transaminase; PT, prothrombin time; HRR, Hemoglobin/RDW;
RLR, RDW/ Lymphocyte count; RPR, RDW/Platelet count; RAR, RDW/ Albumin; PVD, peripheral vascular disease; CVD, cerebrovascular disease; CPD, chronic pulmonary disease; Diabetes Without CC, diabetes without Complications; Diabetes With CC, diabetes with Complications; SLD, severe liver disease; MST, metastatic solid tumor; ACEI, Angiotensin converting enzyme inhibitors; ARB, angiotensin receptor blocker; CCB, Calcium channel blockers.

### 3.2 Exploring the value of RDW-derived indices in predicting 1-year outcomes in patients with AHF

In order to improve the prediction accuracy and weaken the influence of multicollinearity, we applied the Lasso regression method to screen out the key variables for the five RDW-derived coefficients mentioned above. By observing the path diagram of the regression coefficients (see Figure 2), it can be seen that the regression coefficients gradually converge until zero as the value of λ increases. Figure 3 demonstrates the cross-validation plot. In order to incorporate fewer variables while ensuring the fitting effect, we selected the non-zero coefficients corresponding to λ-1se and included them in the subsequent Cox model. Specifically, HRR, RLR, RPR, RAR, and RDW × MCV incorporate 36, 35, 38, 35, and 33 variables, respectively (see supplementary Table 1 for details).

**Figure 2.**
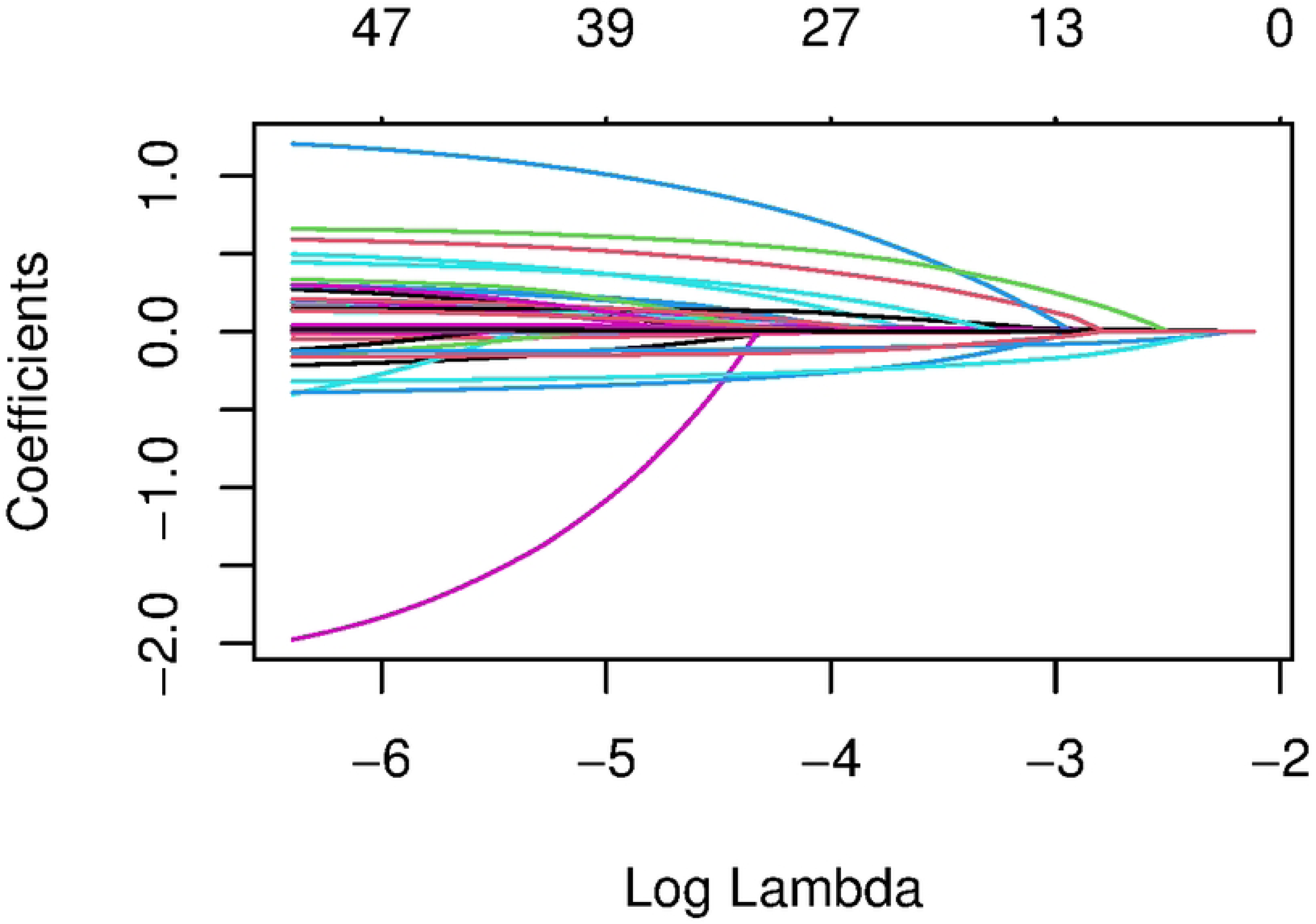
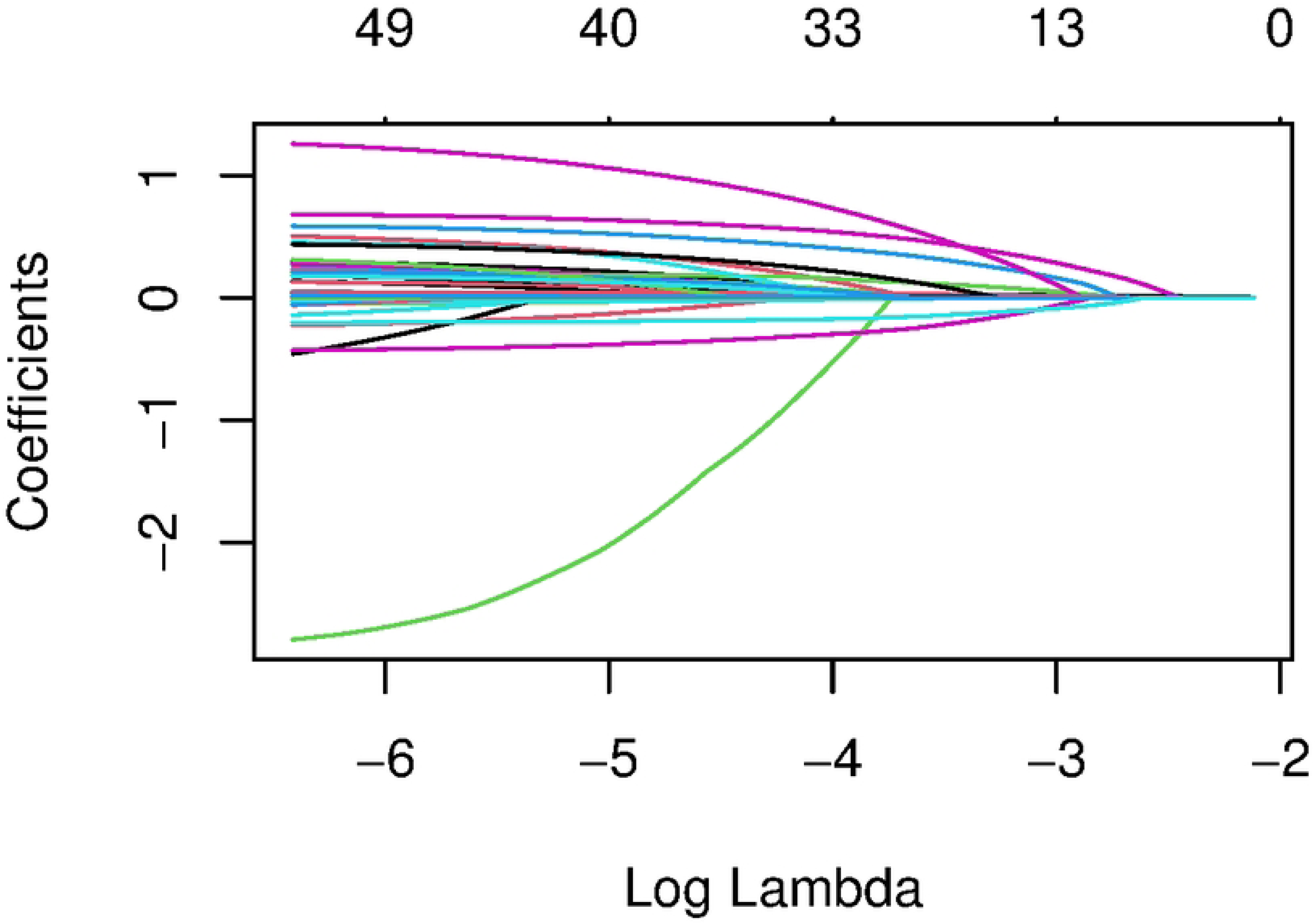
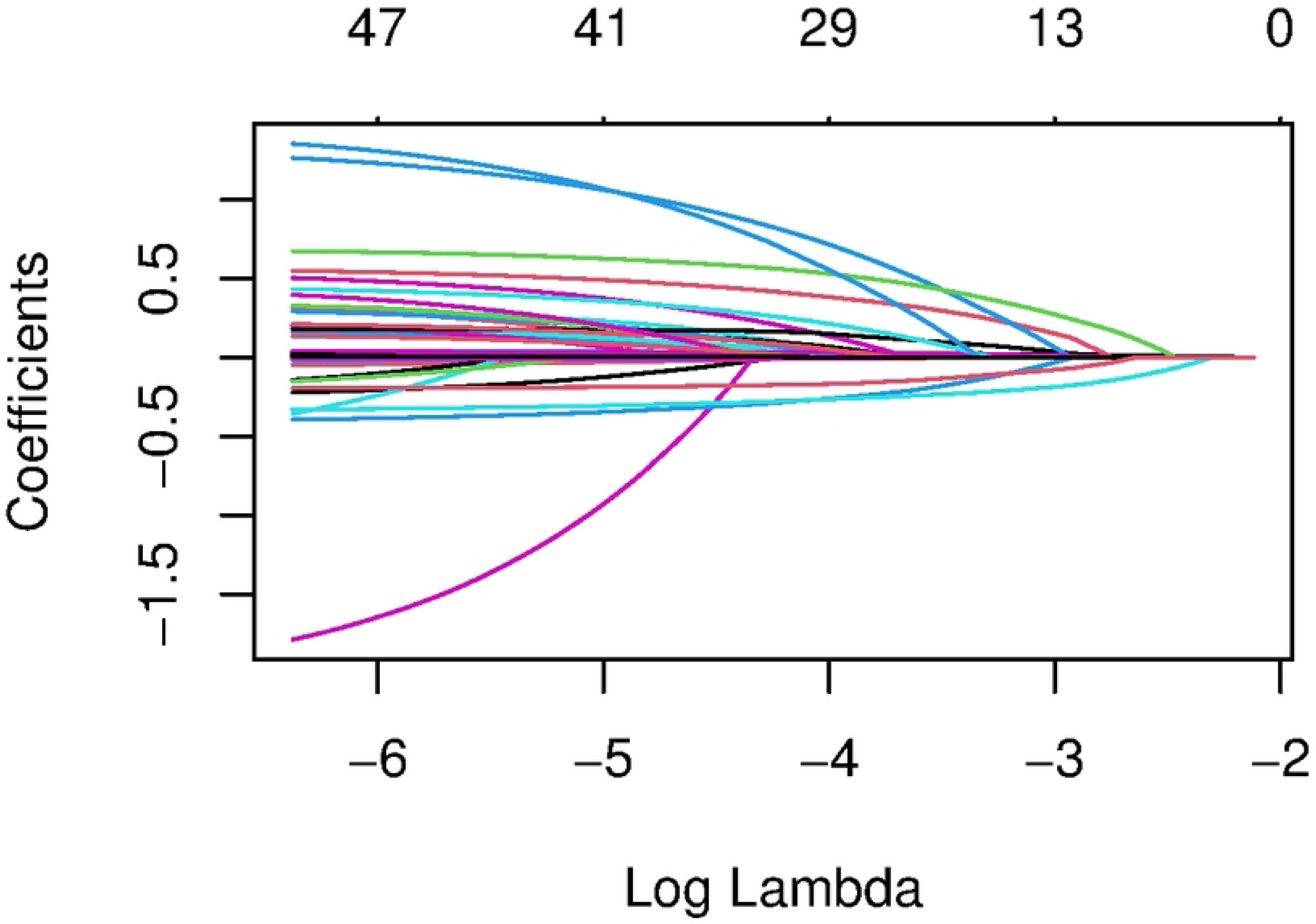
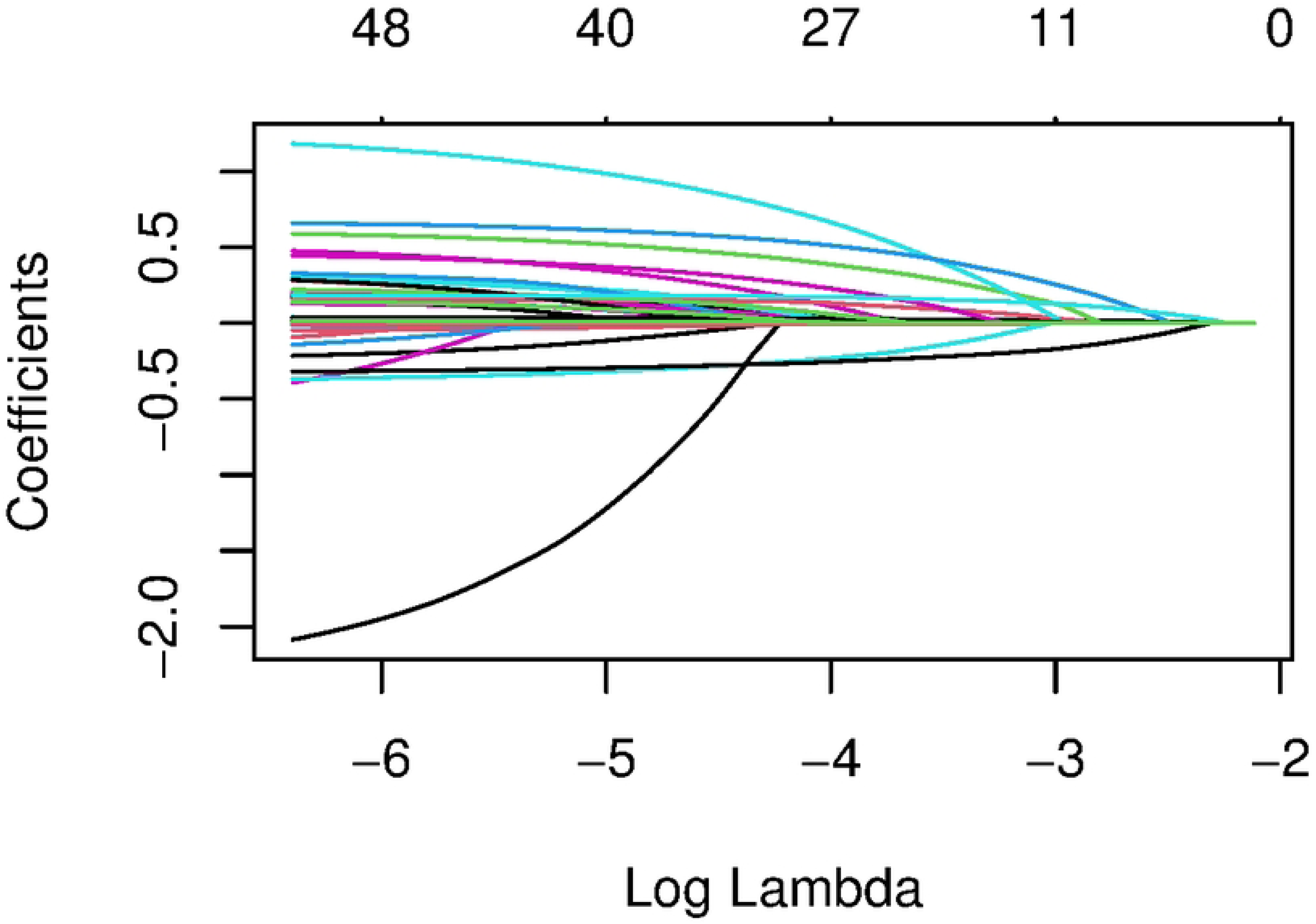
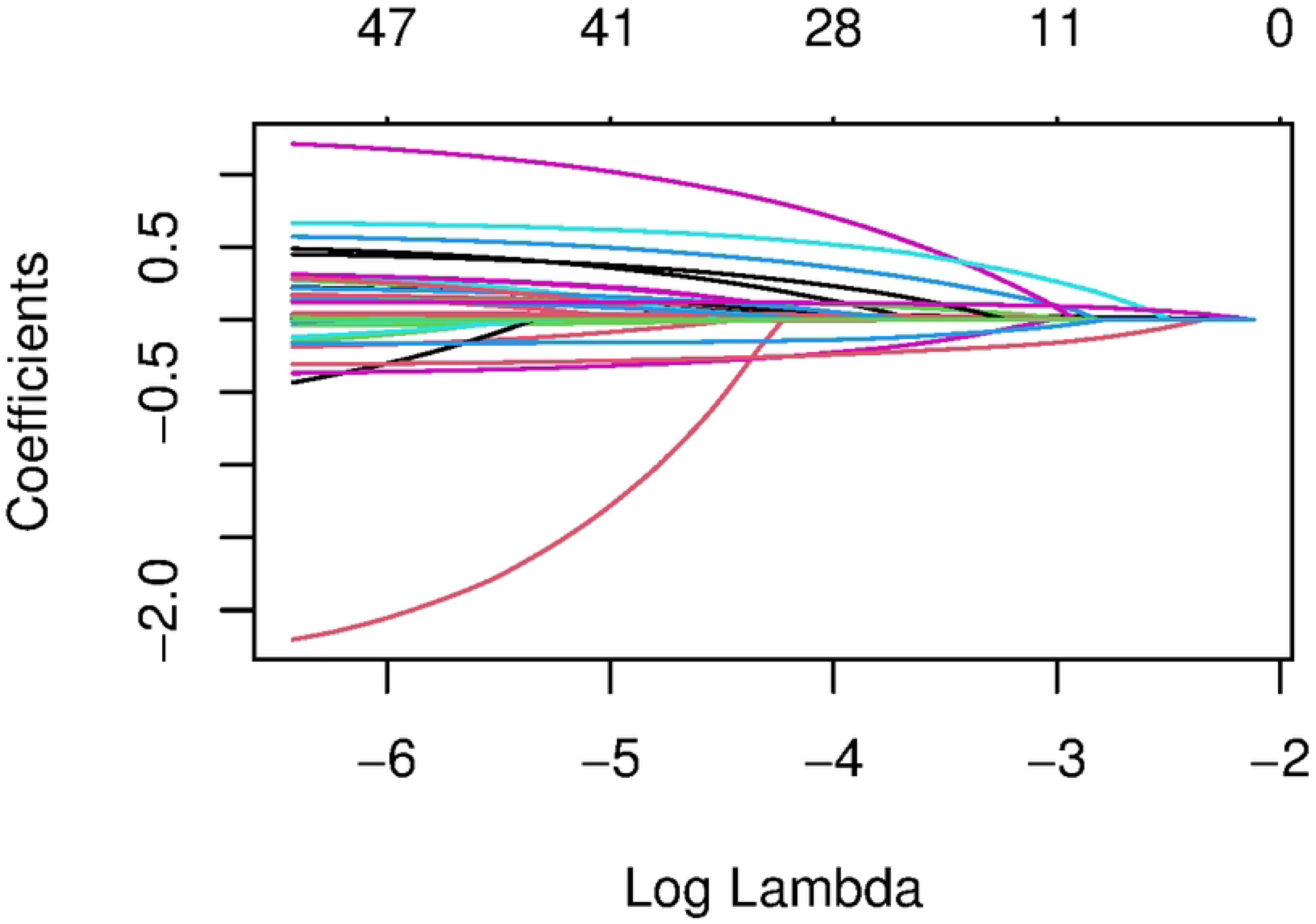
Path diagrams of the respective Lasso regression coefficients of the RDW derivative indices. The horizontal coordinate represents the value of log(λ), the vertical coordinate represents the value of the coefficients, and each curve represents the coefficients of an independent variable as λ changes. The regression coefficients gradually converge until zero as the value of λ increases. From top left to bottom right are HRR, RLR, RPR, RAR, and RDW × MCV.

**Figure 3.**
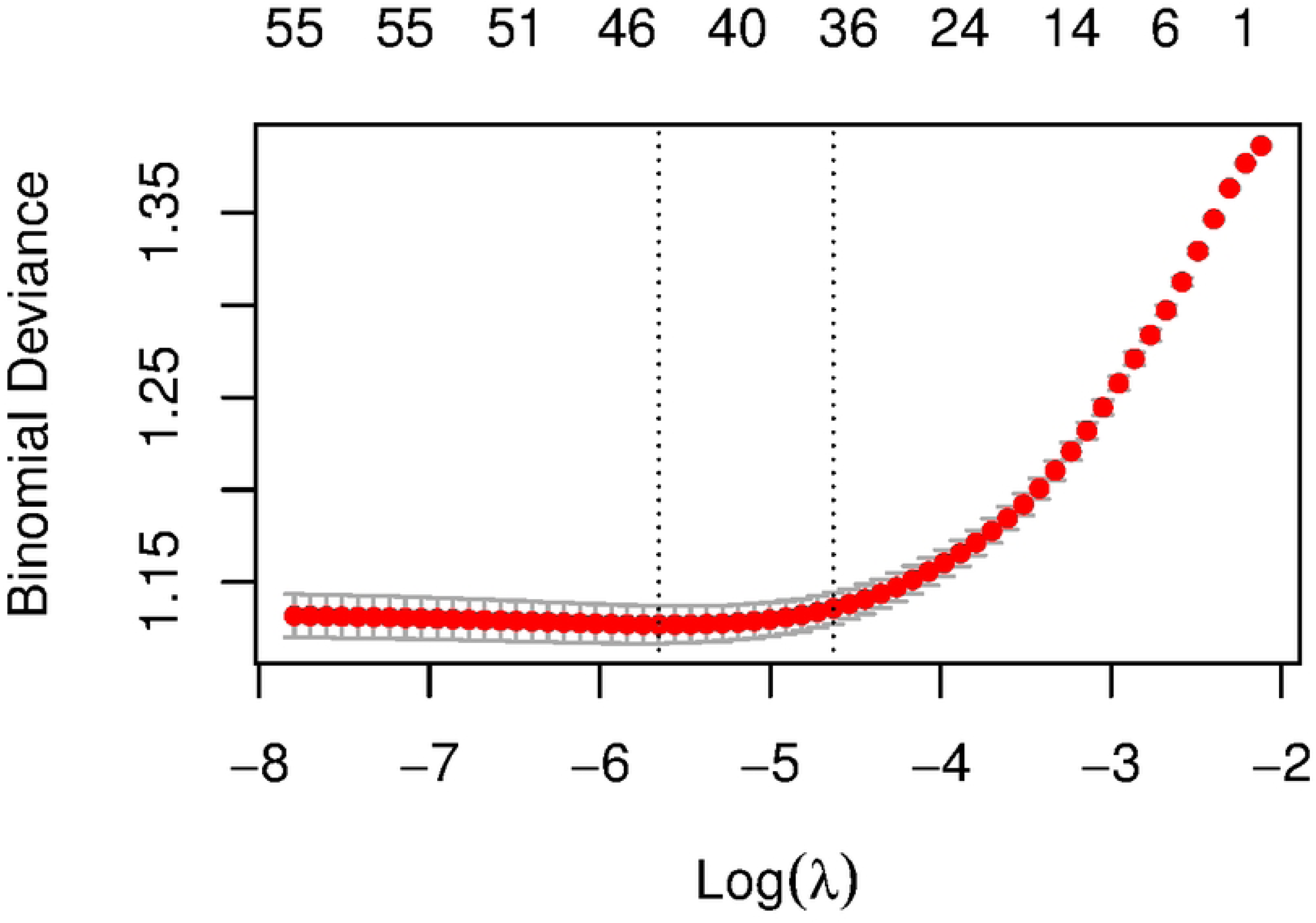
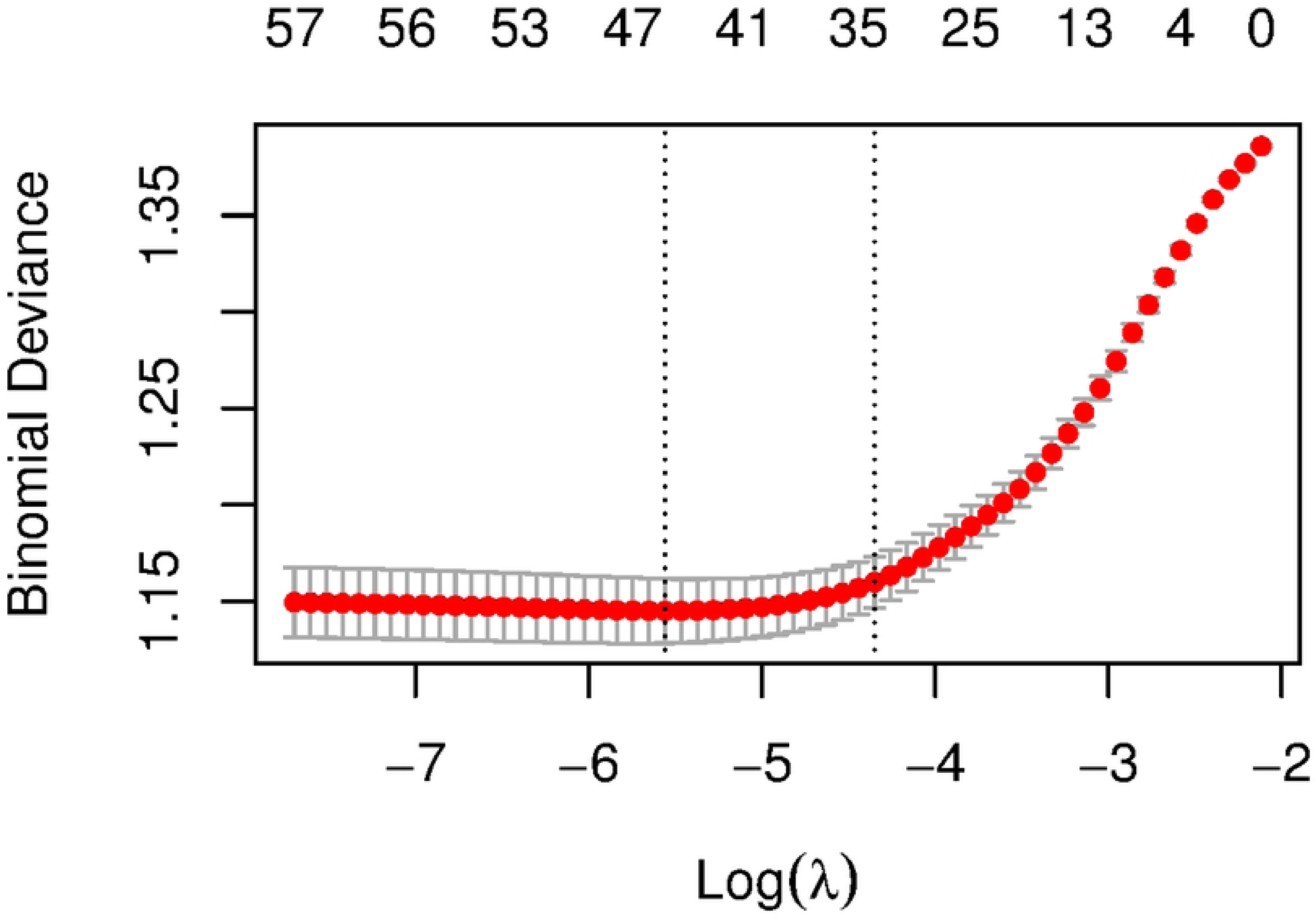
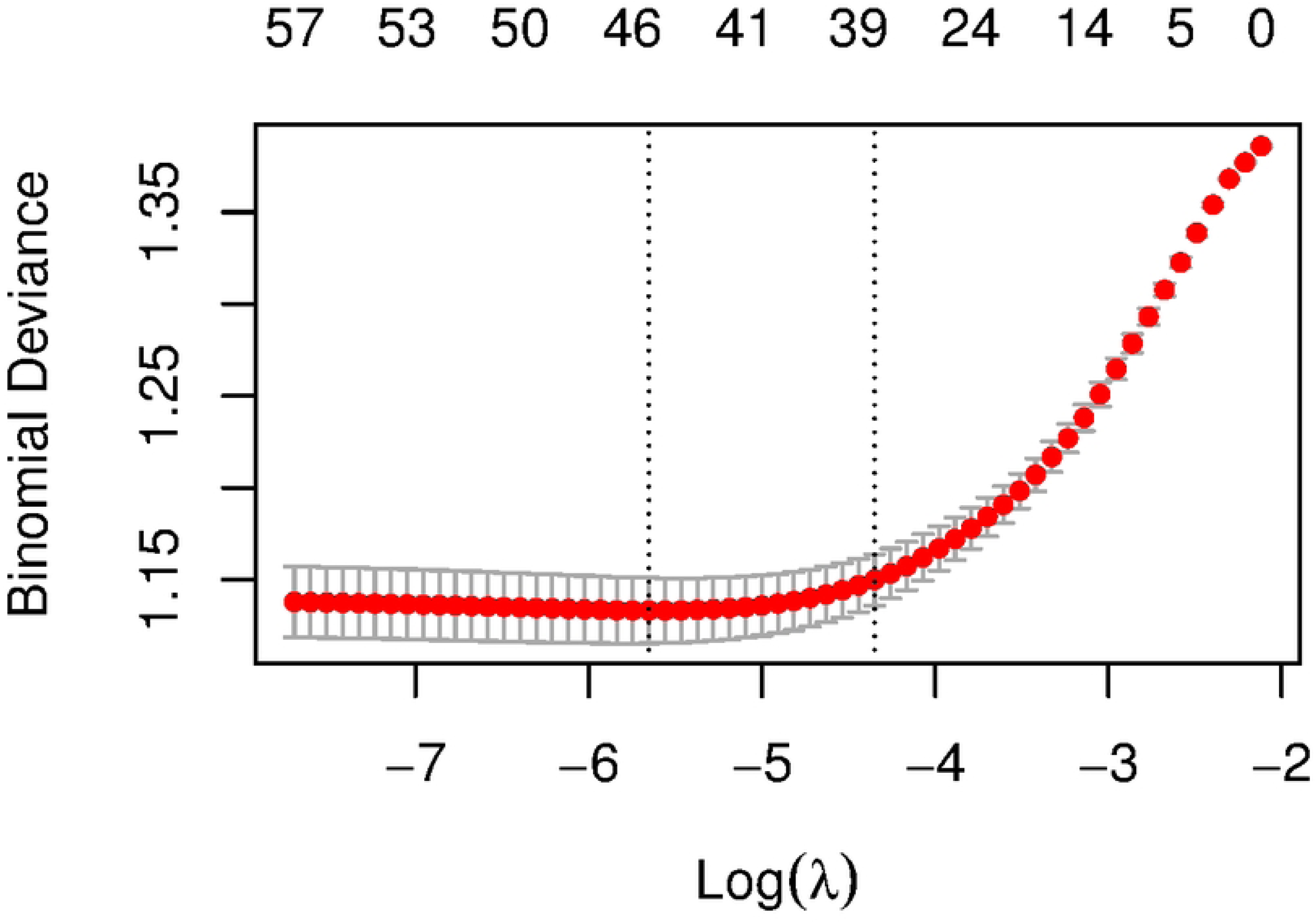
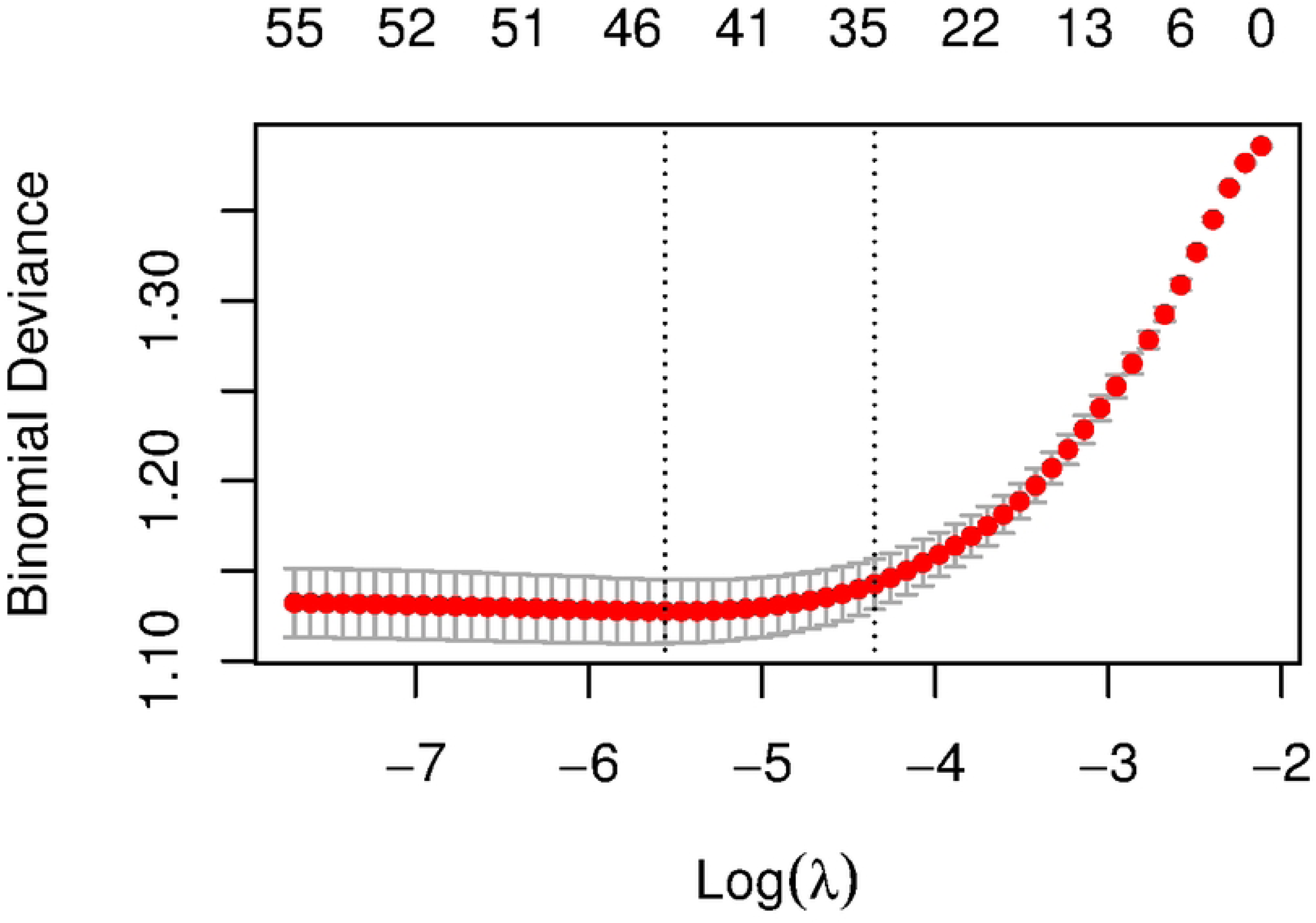
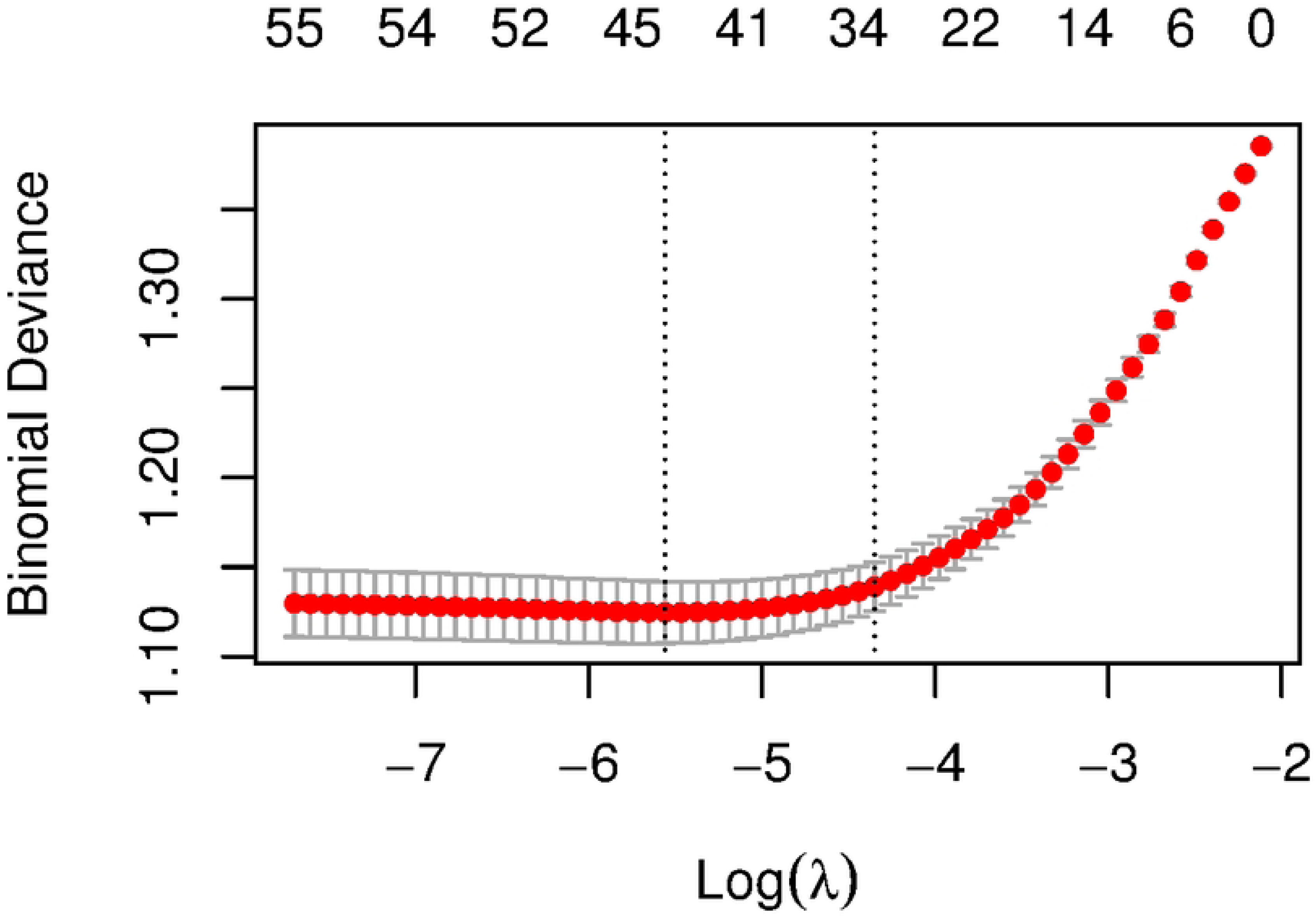
Cross-validation plots of the respective lasso regressions for the RDW derivative indices. Cross-validation curves are used to select the optimal value of λ. The x-axis is the log(λ) of the penalty coefficients, and the y-axis is the deviation of the model. There are usually two dashed lines, λ_min on the left, indicating the λ with the smallest deviation, and λ-1se on the right, indicating the value of λ when the deviation is close to the minimum but the model is more concise. This study hangs on the rightmost λ-1se. From top left to bottom right are HRR, RLR, RPR, RAR, and RDW × MCV. Theirλ-1se is equal to 36, 35, 38, 35, and 33 respectively.

Subsequently, we substituted each of these screened variables into a univariate Cox model, which showed that the p-values of these five RDW correlation coefficients were less than 0.001 (see Table 2). Further, we included the variables with P-values less than 0.05 (see Exhibit 2) in a multivariate Cox model to correct for confounders. The results showed that all four variables, except RLR, were independent predictors in patients with AHF (see Table 2), and RPR had the highest HR (HR = 1.89, 95% CI = 1.33-2.67), which means that the risk of death was increased by 1.89-fold for each unit increase in RPR.

**Table 2.** Univariate and Multivariate Cox proportional risk modeling results for RDW derivative indexes.

| Variables | Univariate analysis |  | Multivariate analysis |  |
| --- | --- | --- | --- | --- |
|  | HR ( 95%CI ) | P value | HR ( 95%CI ) | P value |
| HRR | 0.84 (0.82 ~ 0.86) | <.001 | 0.90 (0.88 ~ 0.93) | <.001 |
| RLR | 1.01 (1.01 ~ 1.01) | <.001 |  |  |
| RPR | 3.22 (2.47 ~ 4.22) | <.001 | 1.89 (1.33 ~ 2.67) | <.001 |
| RAR | 1.26 (1.22 ~ 1.29) | <.001 | 1.12 (1.09 ~ 1.16) | <.001 |
| RDW×MCV | 1.16 (1.14 ~ 1.18) | <.001 | 1.07 (1.05 ~ 1.10) | <.001 |

HRR, hemoglobin-to-RDW ratio; RLR, RDW-to-lymphocyte ratio; RPR, RDW-to-platelet ratio; RAR, RDW-to-albumin ratio; RDW×MCV, the product of RDW and MCV; RDW, red cell distribution width; MCV, mean corpuscular volume. HR, Hazard Ratio.

### 3.3 Assessment of the effect of RDW correlation coefficients on survival in patients with AHF

We used Jamovi software to calculate the optimal cut-off values for the four independent predictors mentioned above (HRR, RPR, RAR, and RDW × MCV) for use in survival analysis (see Table 3, Attached Figure 1), and classified these four indices into two groups, low and high, based on these cut-off values. Based on this, we plotted survival curves (see Fig.4), which showed a gradual decrease in survival for each index over time. Taking the low value group as a reference, the risk of death decreased in the high HRR group (HR=0.560, 95%CI=0.512-0.611), whereas it increased in the high RPR, RAR and RDW×MCV groups (HR = 1.612, 95%CI = 1.441-1.803; HR = 1.821, 95%CI = 1.670-1.985; HR = 1.916, 95%CI = 1.755-2.091).

**Figure 4.**
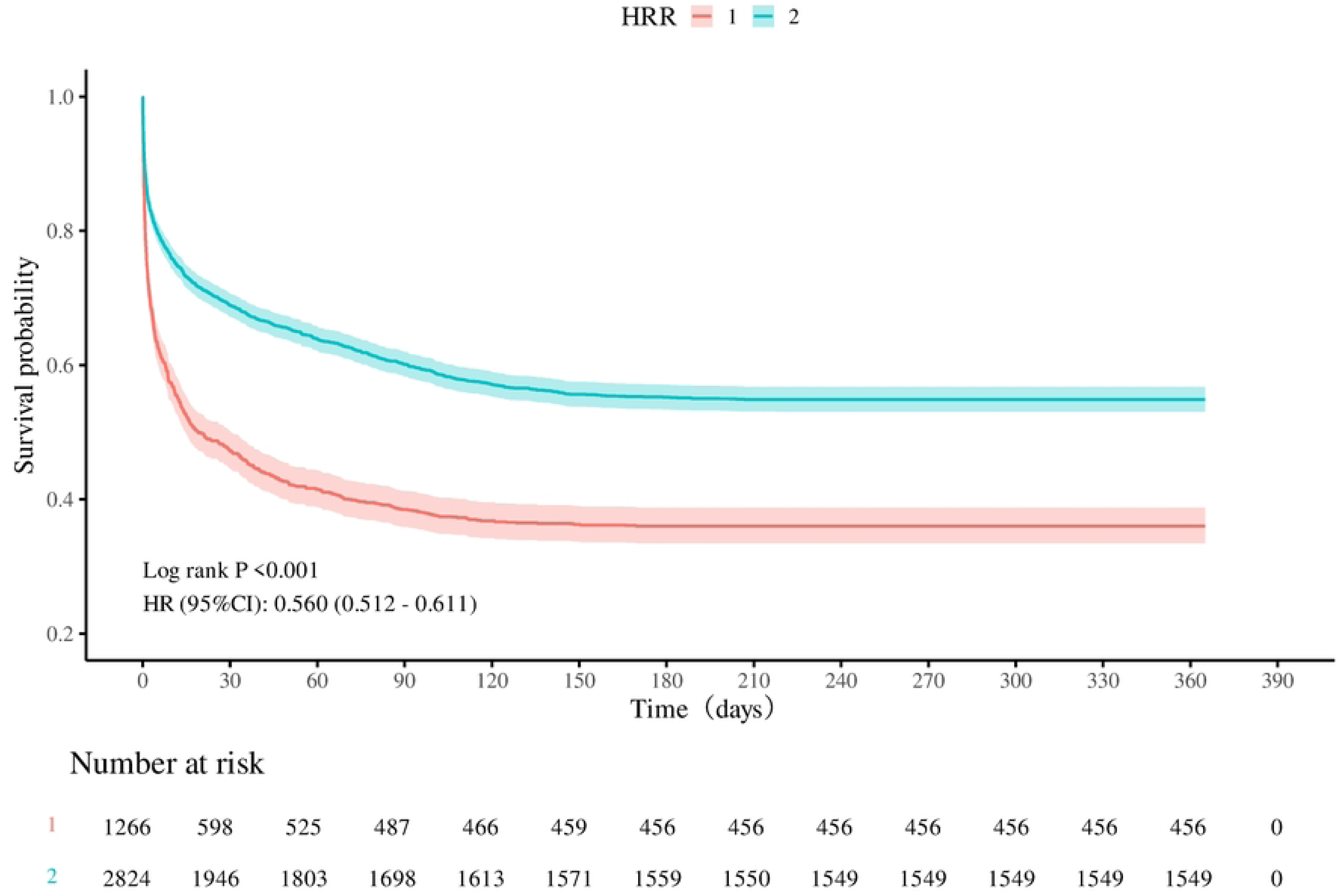
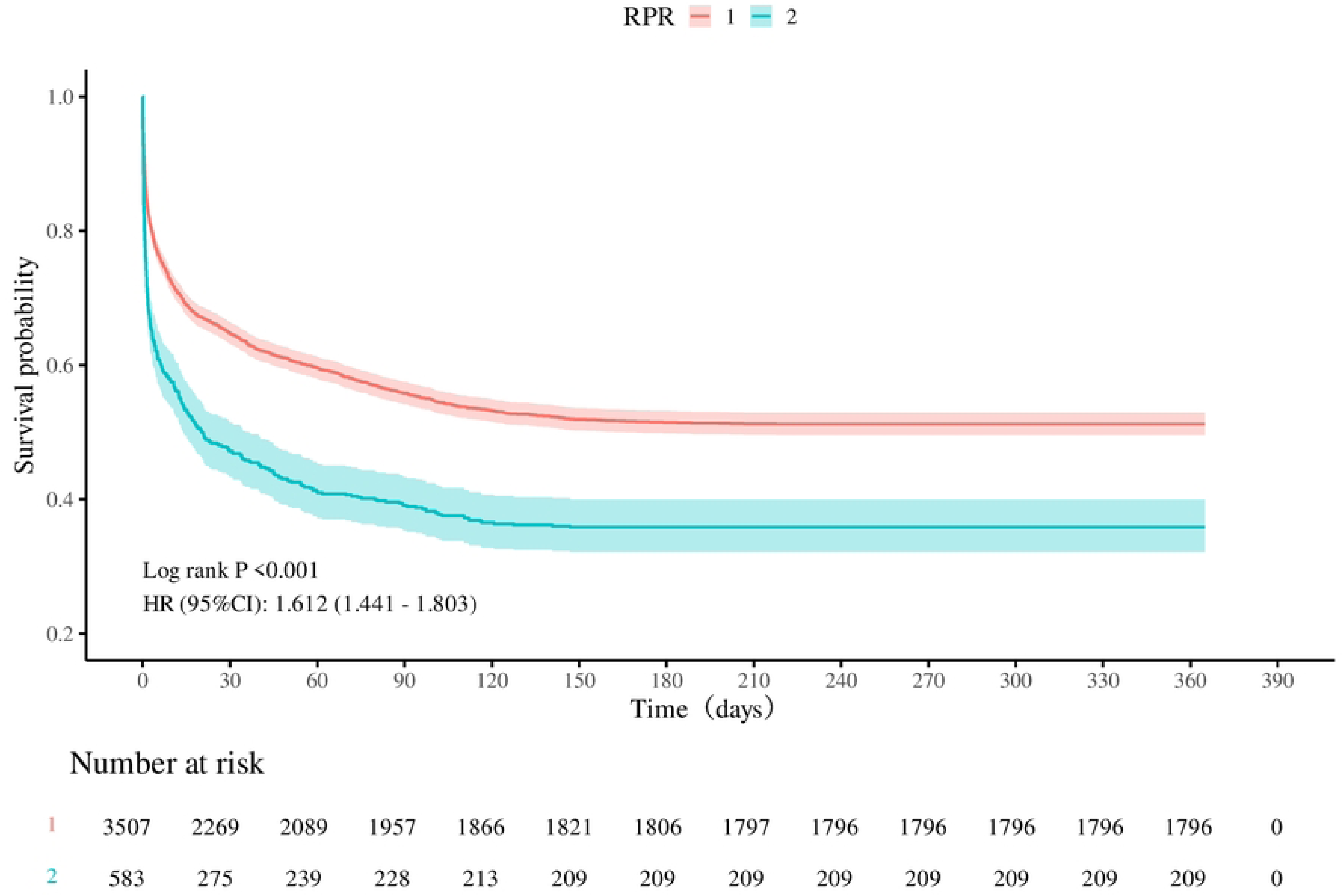
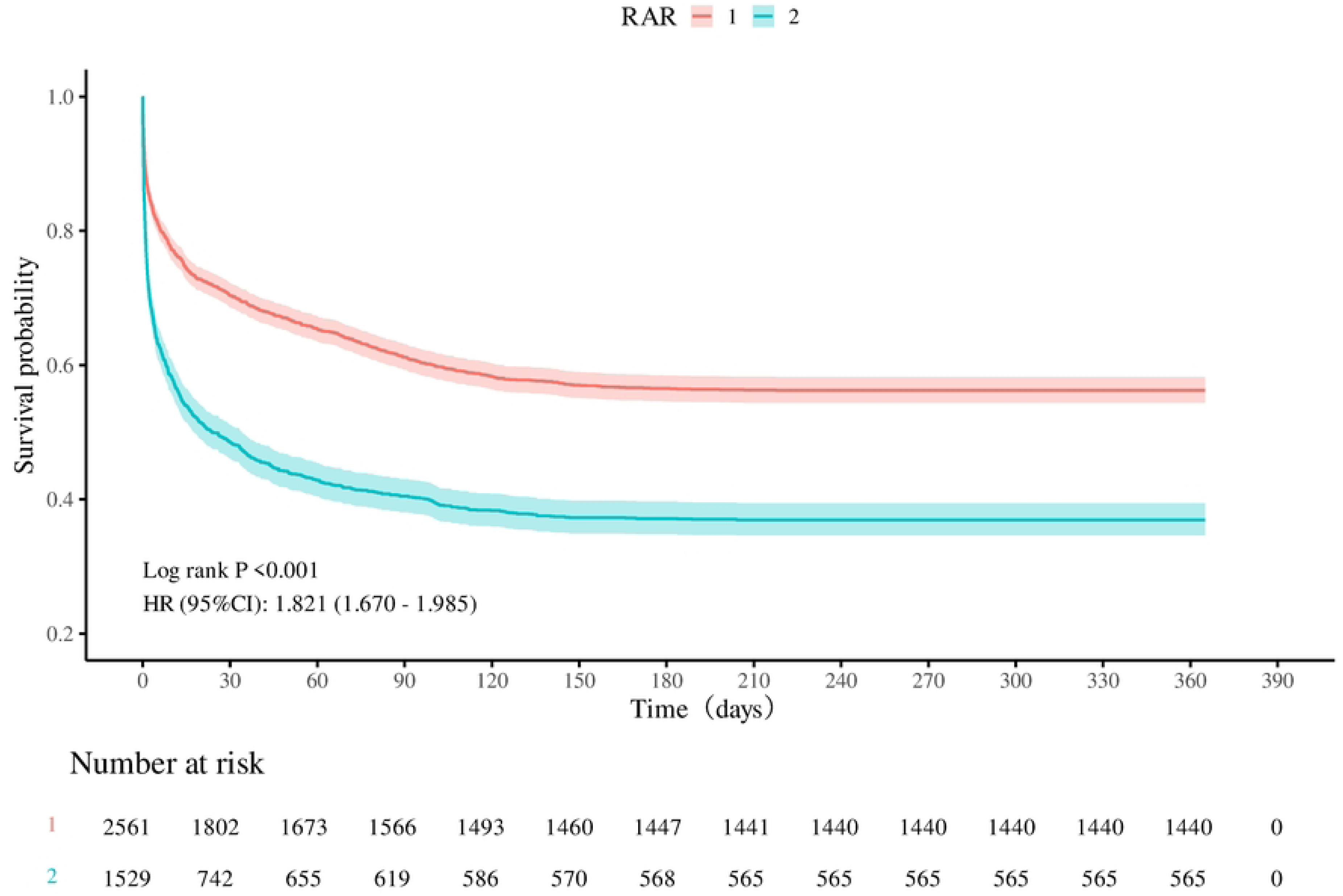
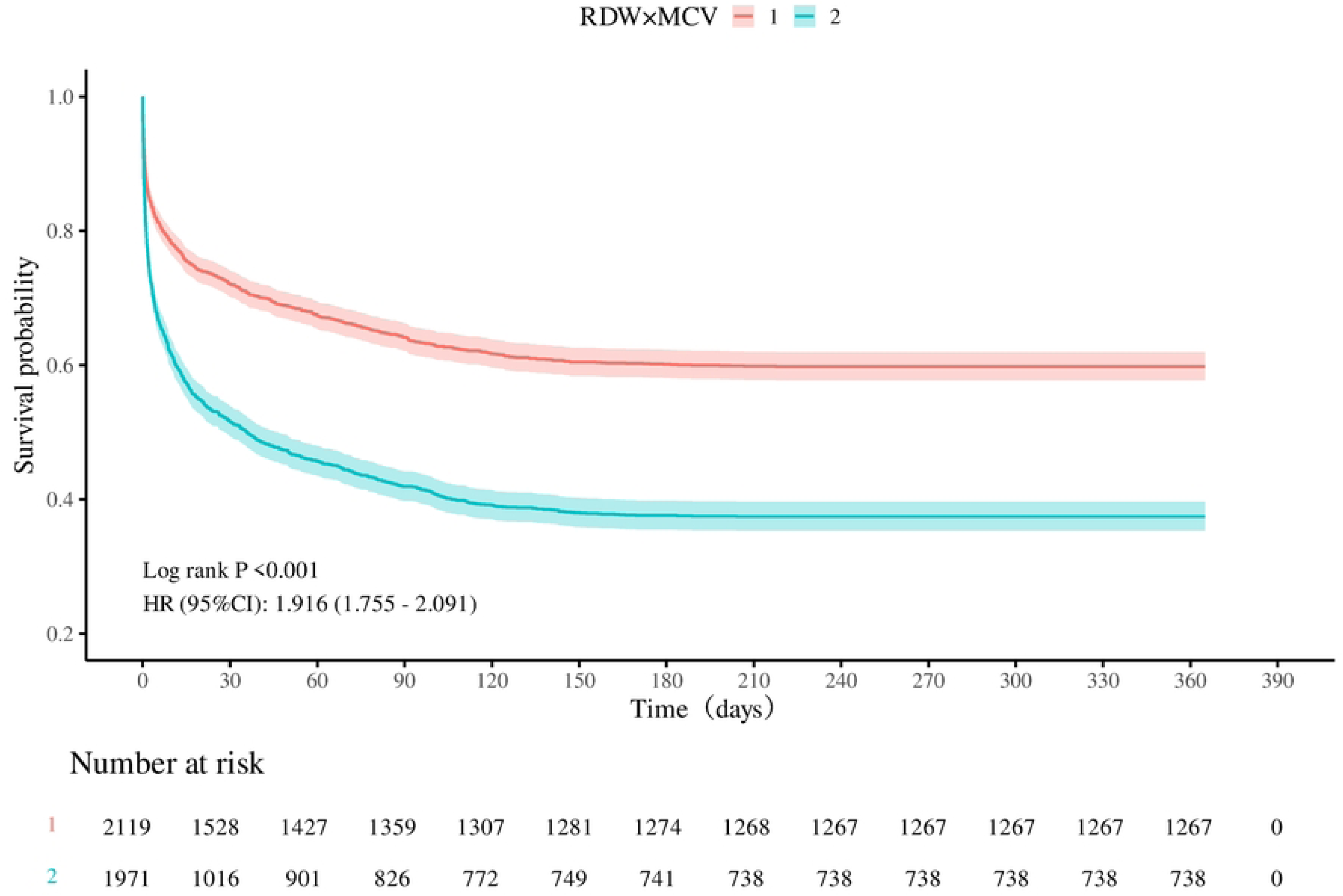
The Kaplan-Meier survival curve for acute heart failure patients within one year of follow-up based on Cox regression. The horizontal axis represents survival time, and the vertical axis represents survival rate. From top to bottom are the survival curves for the RDW-derived indices (HRR, RPR, RAR, RDW × MCV) that have independent predictive value for acute heart failure.

**Table 3.** Cut-off values for each RDW-derived index.

|  | HRR | RPR | RAR | RDW×MCV |
| --- | --- | --- | --- | --- |
| Cut-off | 5.540 | 0.13 | 4.85 | 14.11 |

HRR, hemoglobin-to-RDW ratio; RLR, RDW-to-lymphocyte ratio; RPR, RDW-to-platelet ratio; RAR, RDW-to-albumin ratio; RDW×MCV, the product of RDW and MCV; RDW, red cell distribution width; MCV, mean corpuscular volume.

### 3.4 Assessment of the efficacy of RDW-derived indices in predicting 1-year outcomes in patients with AHF

To assess the predictive efficacy of each index for the risk of death in AHF, we plotted time-ROC curves at 30 days, 90 days, and 1 year (see Figure 5). The results showed that RDW×MCV had the largest area under the curve, i.e., it had the highest predictive efficacy (AUC = 0.612-0.613).

**Figure 5.**
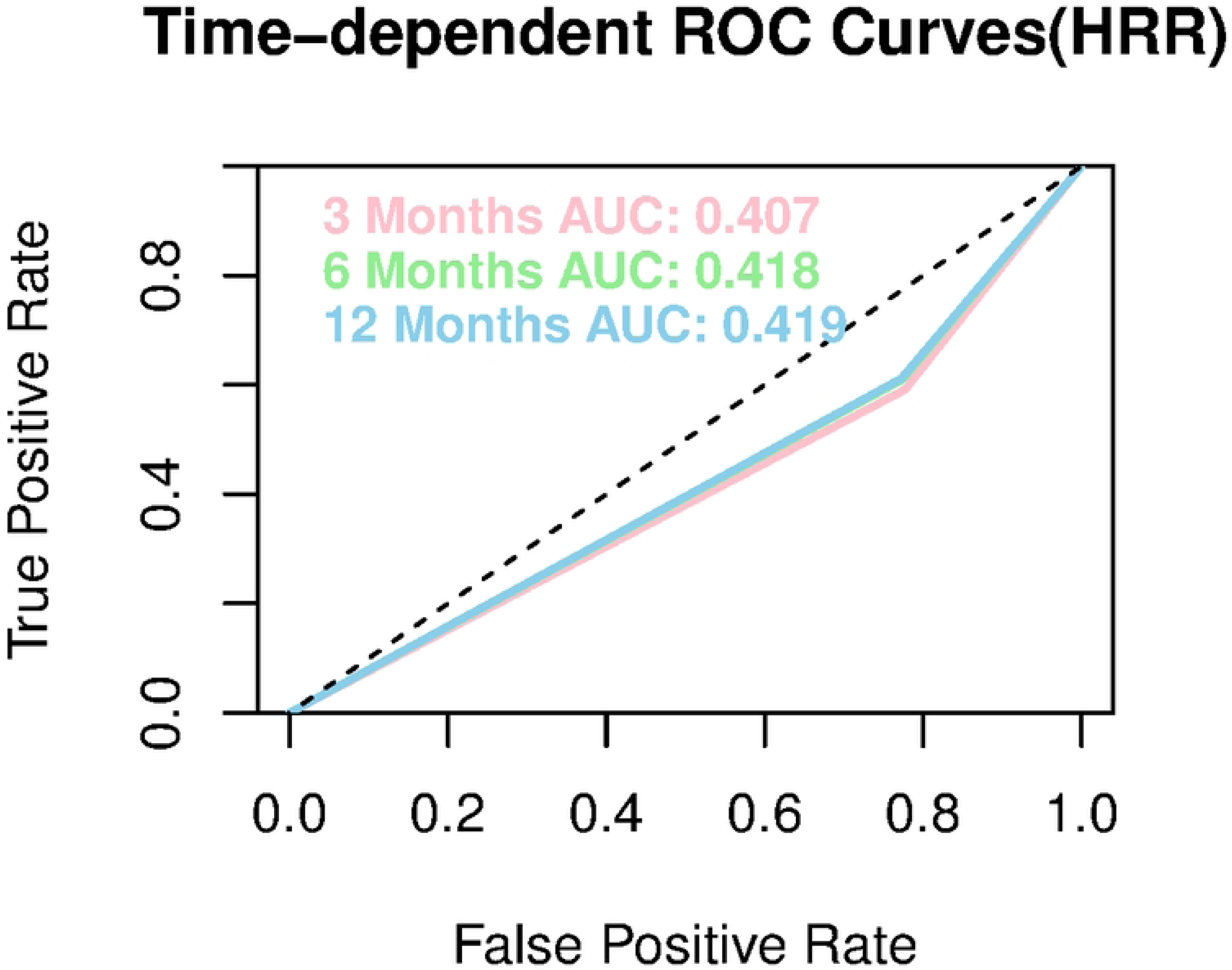
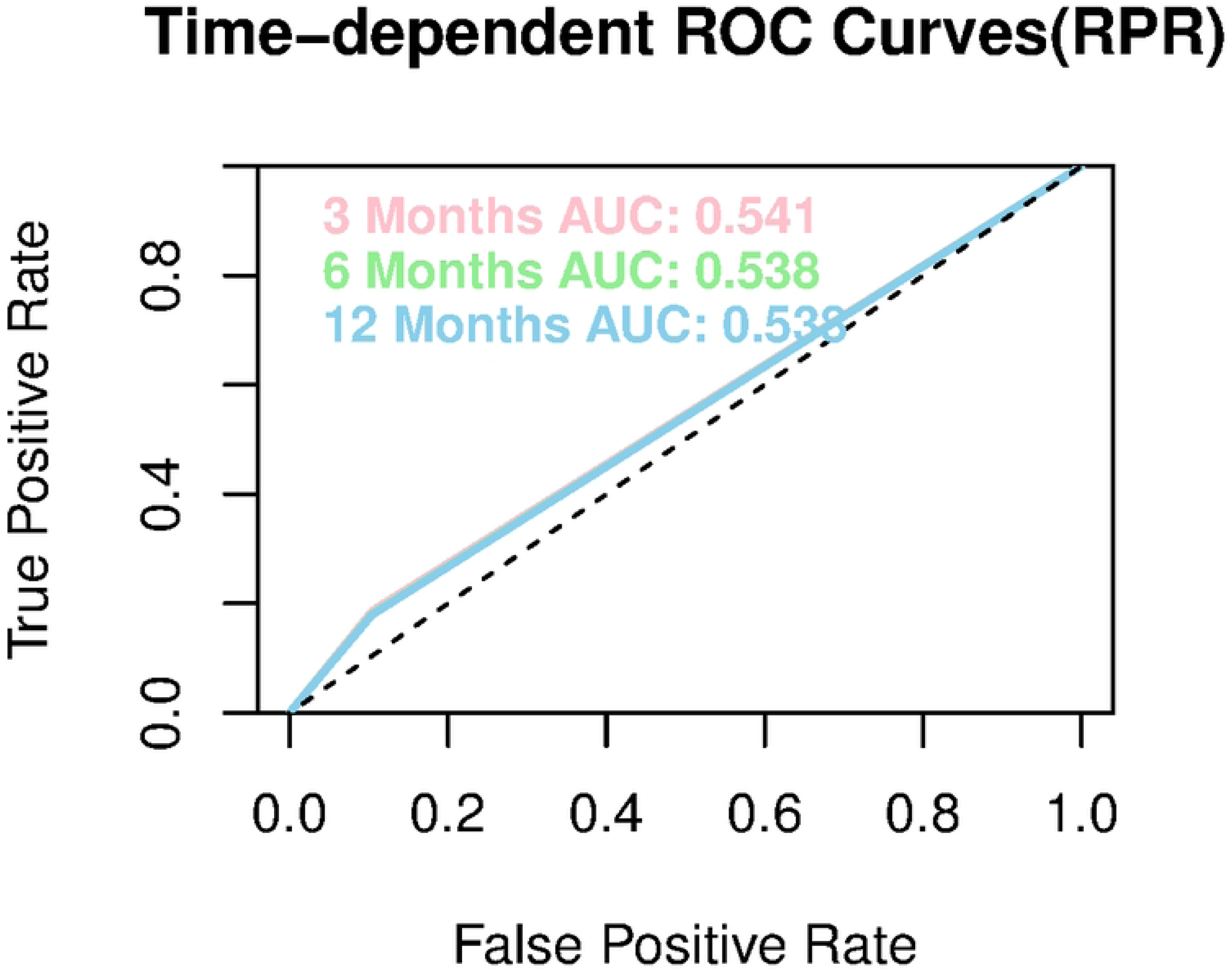
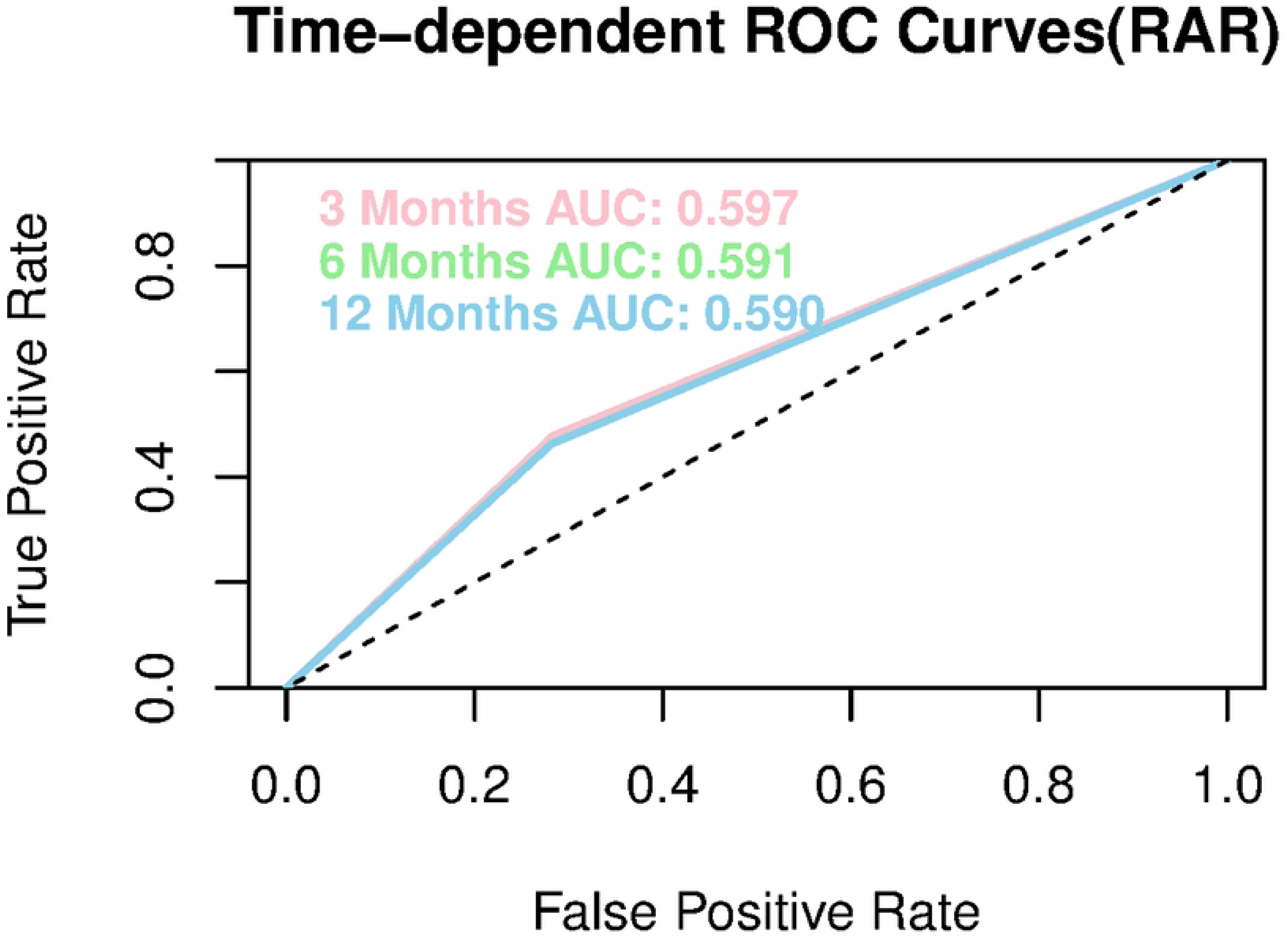
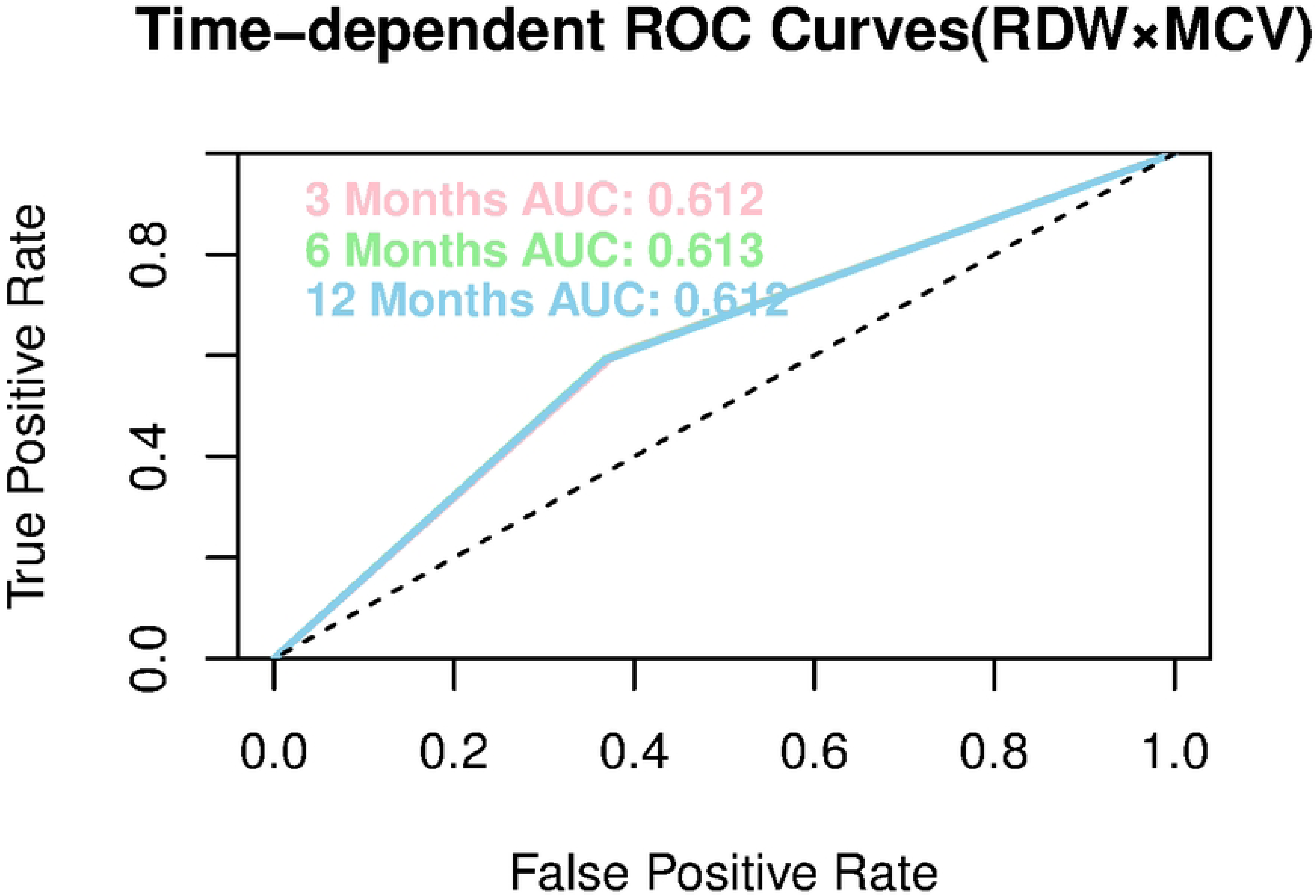
Analysis of ROC curves over time for RDW-derived indices with independent predictive value for acute heart failure. The horizontal axis indicates the false-positive rate, which can also be referred to as 1-specificity; the vertical axis indicates the true-positive rate. To describe the predictive performance of the RDW-derived index for poor prognosis in acute heart failure over time. Pink: three months; green: six months; blue: 12 months.

### 3.5 Exploring nonlinear associations between RDW-derived indicators and outcomes

To explore whether there was a nonlinear association between the above variables and the 1-year mortality outcome, we included confounding variables screened in supplementary Table 2 and plotted RCS (see Figure 6) after adjusting for covariates. This study found a statistically significant relationship between HRR, RPR, RAR, and RDW×MCV and outcome (P for overall < 0.001), but this relationship was more likely to be linear than nonlinear (P for nonlinear > 0.05).

**Figure 6.**
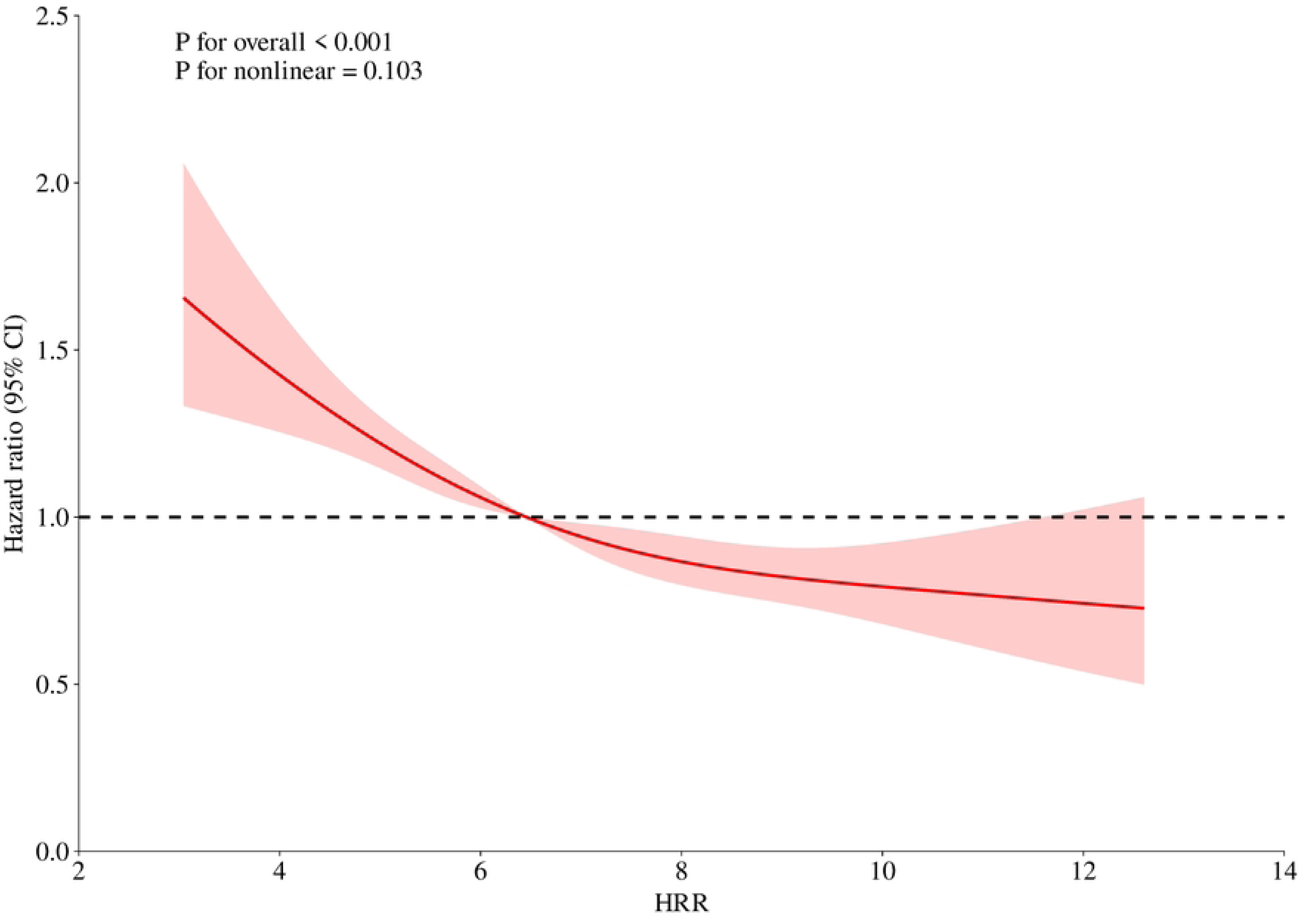
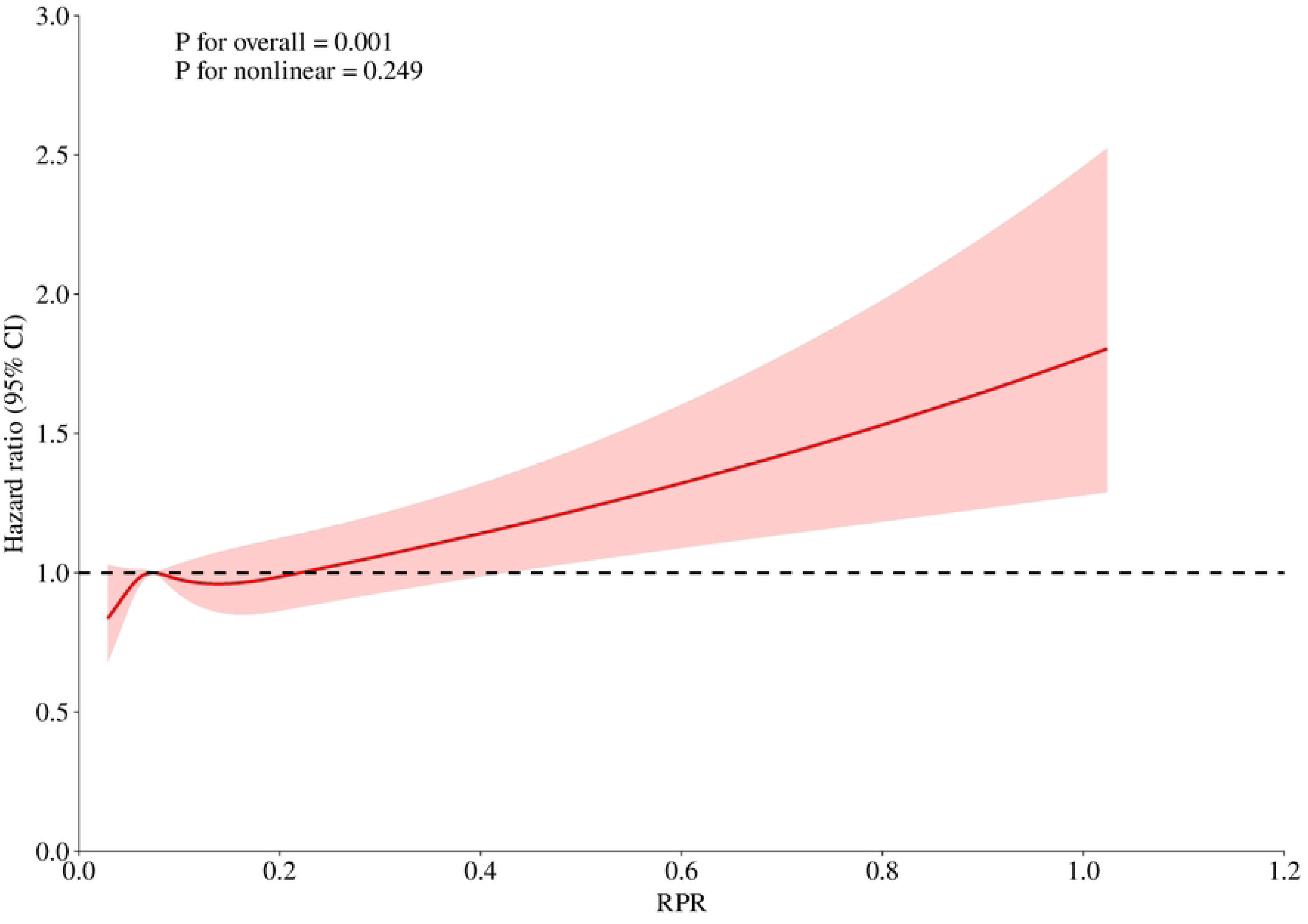
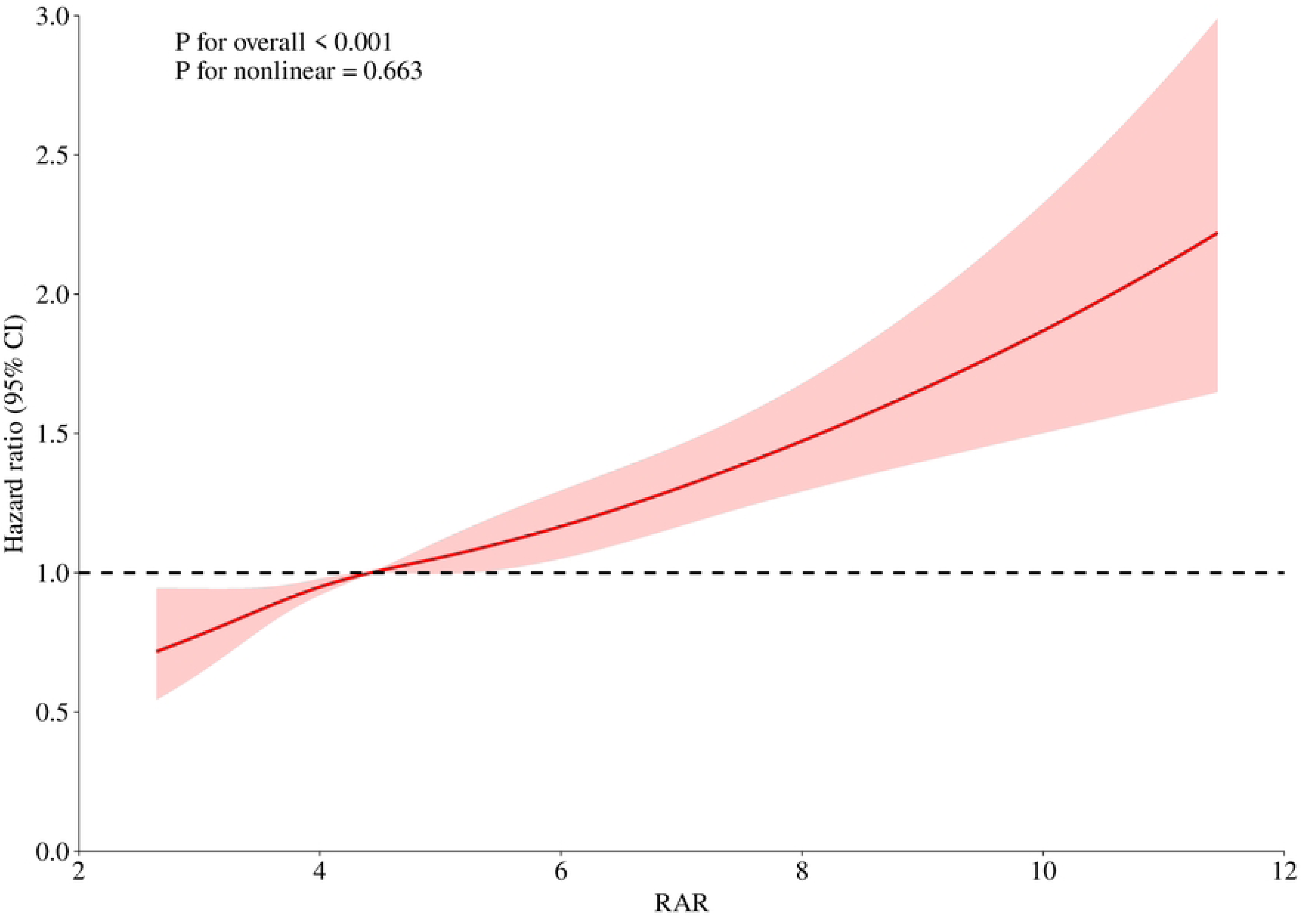
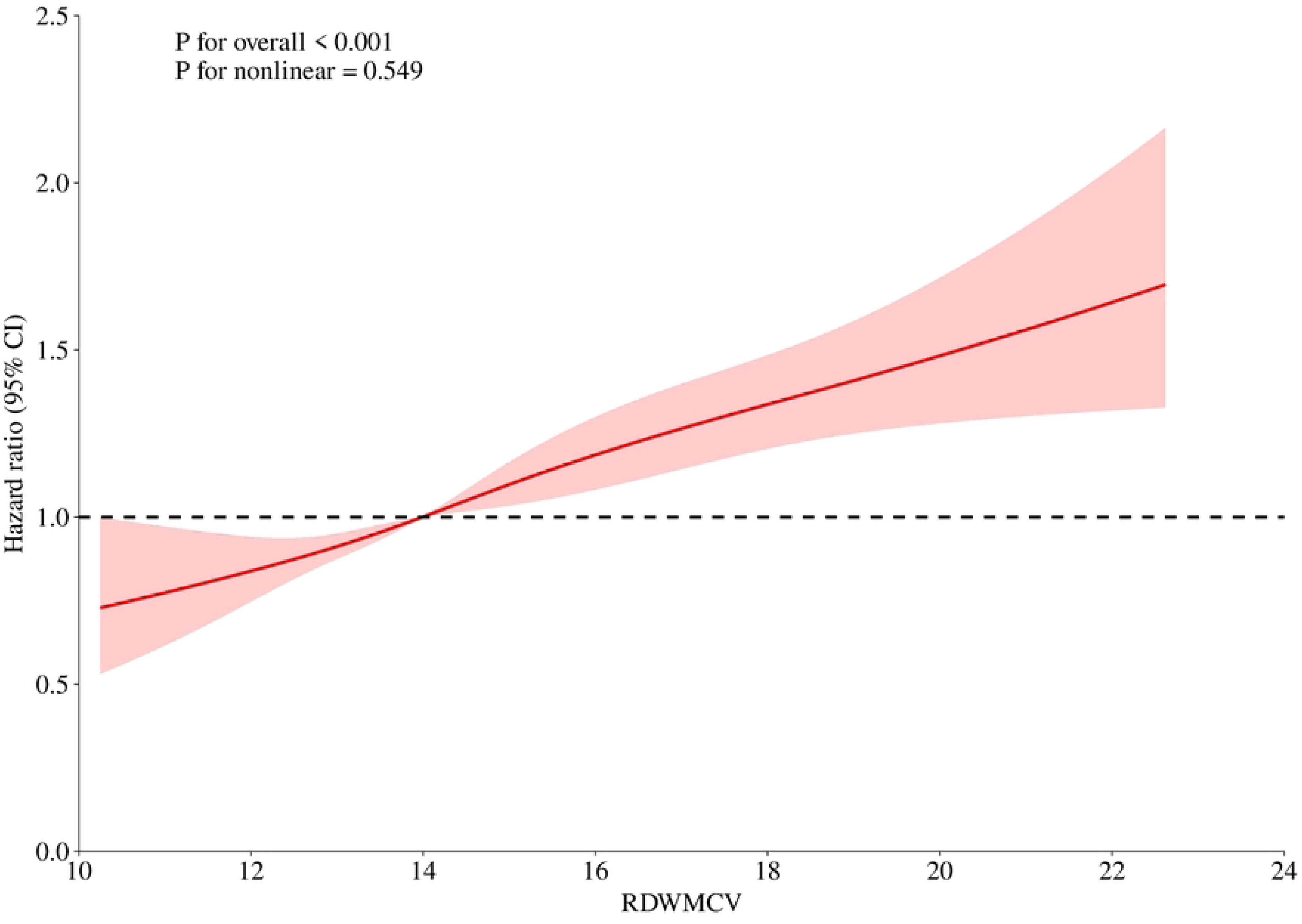
Restricted cubic lines for RDW derivative indexes adjusted for covariates Attached Figure 1 HRR, RPR, RAR, and RDW × MCV cut-offs.

## 4 Discussion

In this study, we found that the RDW-derived coefficients HRR, RPR, RAR, and RDW × MCV were effective in predicting 1-year mortality in patients with AHF and were independent of other prognostic markers of AHF (e.g., SBP, HR, and NT-proBNP) (20, 21), with the strongest effect on survival time being that of RPR (HR = 1.89, 95% CI = 1.33-2.67), and the time-ROC results showed the highest predictive efficacy of RDW × MCV, for which the independent predictive value of RLR has not been found. And we found that the relationship between HRR and outcome was negative, whereas RPR, RAR, and RDW × MCV were positive, whereas this relationship was more likely to be linear.

### 4.1 HRR and AHF

The association between HRR and AHF has been investigated. A prospective study with a median follow-up of 3.1 years showed that lower HRR was associated with an increased risk of death in patients with ADHF(14). Furthermore, in elderly patients with AHF with preserved ejection fraction, HRR was an independent risk factor for the prognosis of major short-term adverse cardiovascular events and was negatively associated with the occurrence of these events(22). Consistent with previous studies, we found that low HRR was associated with poor prognosis in AHF and further confirmed its use as an independent predictor of 1-year mortality in patients with AHF.

### 4.2 RPR and AHF

Most previous studies have focused on the association between RPR and acute myocardial infarction (23, 24), while few studies have explored its potential value in HF. A small study of canines with HF due to mucinous neoplastic mitral valve disease showed a reduction in RPR in dogs that were symptomatic and poorly responsive to treatment or presented with refractory chronic HF compared to healthy dogs, suggesting that this parameter may be an aid in assessing disease severity(16). In this study, we investigated for the first time the relationship between RPR and 1-year prognosis in patients with AHF and found that RPR tended to be elevated in patients with a poor prognosis of AHF and that there was a significantly strong association between RPR and the risk of death within 1 year, after adjusting for potential confounders.

### 4.3 RAR and AHF

Current studies have focused on the association between RAR and prognosis in cardiovascular disease, with RAR demonstrating potential as a biomarker for prognostic assessment in patients with acute myocardial infarction and atrial fibrillation, with a significantly increased risk of death, particularly in patients with higher RAR levels(17, 25, 26). In addition, the present study further reveals that RAR may also be a candidate marker for prognostic assessment of AHF, finding that high RAR levels are associated with a higher risk of death in this patient population.

### 4.4 RDW × MCV and AHF

One study noted that the addition of MCV enhances the prognostic validity of RDW as a predictor of overall survival(27). Another study showed that the inclusion of MCV in the consideration significantly enhanced the association between RDW and hypertension-induced target organ damage(28). In the present study, we pioneered the relationship between the product of RDW and MCV and AHF and found that it not only predicted poor prognosis of AHF as an independent factor, but also demonstrated the highest predictive efficacy.

### 4.5 RLR and AHF

Elevated RDW with decreased relative lymphocyte count has been identified as an independent predictor of death in patients with advanced HF(29), however, this paper did not obtain a positive result for RLR in predicting AHF, which may be due to the inclusion of the variables as well as the different study populations.

### 4.6 Potential mechanisms of the relationship between RDW-derived indicators and AHF

Inflammation seems to play a crucial role when discussing the relationship between RDW-derived indices and AHF. Inflammatory mediators such as C-reactive proteins, TNF-α, and IL-6 contribute to the development of AHF(30)and damage the bone marrow along with premature erythrocyte entry into the bloodstream(31). Among them, TNF-α and IL-6 inhibit the proliferation and differentiation of bone marrow erythroid progenitor cells and reduce renal erythropoietin production(32), resulting in a decrease in Hb and an increase in the RDW, while bone marrow dysfunction can also lead to an increase in MCV(33). During the inflammatory response, vascular endothelial cells are damaged and activate platelets, resulting in excessive platelet depletion and destruction, leading to a decrease in PLT in the peripheral blood(34). In addition, inflammation increases capillary permeability, leading to leakage of plasma components (e.g., albumin) through the capillaries into the interstitium(35).

Previous studies have shown that elevated oxidative stress is associated with worsening of AHF(36). In addition to inflammatory factors, oxidative stress may be another key factor contributing to changes in HRR, RAR, and RDW × MCV in patients with AHF (33, 37, 38). Notably, malnutrition is quite common in patients with AHF and is associated with higher 1-year mortality, suggesting that it may also be an important mechanism influencing the prognosis of HF(33, 37–39). However, the mechanism of how the above indicators specifically affect the prognosis of HF is still not fully understood, and more in-depth basic studies are expected to reveal this complex process in the future.

### 4.7 Limitations

Because the data were derived from the MIMIC database, we lacked key indicators such as left ventricular ejection fraction to comprehensively assess the prognosis of AHF. In the future, we plan to incorporate more variables to more accurately evaluate the prognostic significance of RDW-related metrics. In addition, the predictive efficacy of the current RDW-derived indices for AHF is moderate, and we expect to enhance the predictive power of the model by adding more variables to construct a column chart.

## 5 Conclusion

The RDW-derived indices of HRR, RPR, RAR, and RDW × MCV were able to effectively predict mortality within 1 year in patients with acute heart failure as predictors alone, whereas RLR did not demonstrate independent predictive value.

## Author Contributions

Dr. Hongyan Dai, as the project leader, developed the study plan and was responsible for the completeness and accuracy of the data analysis. Miao Zhang played a key role in experimental design and data collection, and participated in the initial writing of the paper. Dr. Jing Zhu was responsible for the text writing, while Dr. Degang Mo was responsible for the data analysis. Drs. Shanshan Yuan and Fanhui Lin ensured the accuracy and completeness of the data and also provided in-depth interpretation of the data analysis results. All authors reviewed and approved the revised final draft.

## Ethics statement

Not applicable

## Data availability statement

The datasets generated and analyzed in this study are available for request and use from the corresponding authors.

## Acknowledgments

Not applicable.

